# Identification of Novel Reproducible Combinatorial Genetic Risk Factors for Myalgic Encephalomyelitis in the DecodeME Patient Cohort and Commonalities with Long COVID

**DOI:** 10.64898/2025.12.01.25341362

**Authors:** JM Sardell, S Das, M Pearson, D Kolobkov, AR Malinowski, LM Fullwood, M Sanna, H Baxter, K McLellan, M Natt, D Lamirel, S Chowdhury, MA Strivens, S Gardner

## Abstract

**Background:** Myalgic encephalomyelitis (also known as ME/CFS or simply ME) has severely impacted the lives of tens of millions of people globally, but the disease currently has no accurate diagnostic tools or effective treatments. Identifying the biological causes of ME has proven challenging due to its wide range of symptoms and affected organs, and the lack of reproducible genetic associations across ME populations. This has prolonged misunderstanding, lack of awareness, and denial of the disease, further harming patients.

**Methods:** We used the PrecisionLife® combinatorial analytics platform to identify disease signatures (i.e., combinations of 1-4 SNP-genotypes) that are significantly enriched in two cohorts of ME participants from DecodeME relative to controls from UK Biobank (UKB). We tested whether the number of these signatures possessed by an individual is significantly associated with increased prevalence of ME in a third disjoint cohort of DecodeME participants. We characterized a number of drug repurposing opportunities for a set of candidate core genes whose disease signatures had the strongest association with ME and which were linked to different mechanisms. We then tested gene overlap between the ME signatures identified and previous studies in long COVID, using two independent approaches to explore these shared genetic commonalities.

**Results:** We identified 22,411 reproducible disease signatures, comprising combinations of 7,555 unique SNPs, that are consistently associated with increased prevalence of ME in three disjoint patient cohorts. The count of reproducible signatures was significantly associated with increased prevalence of ME (*p* = 4×10^-21^), and participants with a top 10% signature count had an odds ratio of disease 1.64 times greater than participants with a bottom 10% signature count, confirming that these genetic signatures increase susceptibility for developing ME.

These disease signatures map to 2,311 genes. We identified substantial overlap between the genes found by this combinatorial analysis and previous studies. We found that the 259 candidate core genes most strongly associated with ME are enriched in disease mechanisms including neurological dysregulation, inflammation, cellular stress responses and calcium signaling. We demonstrated that 76 out of 180 genes previously linked to long COVID in UKB and the US All of Us cohorts are also significantly associated with ME in the DecodeME cohort. These findings allowed identification of many existing and novel repurposing opportunities, including candidates linked to several genes with shared etiology for long COVID.

**Conclusion:** These findings provide further evidence that ME is a complex multisystemic condition where the risk of developing the disease has a very clear genetic and biological basis. They give a substantially deeper level of insight into the genetic risk factors and mechanisms involved in ME. The discovery of so many multiply reproducible genetic associations implies that ME is highly polygenic, which has important consequences for its future study and the delivery of clinical care to patients.

The striking overlap in genes and mechanisms between long COVID and ME (76 / 180 long COVID genes tested) suggests the potential for development of novel or repurposed drug therapies that could be used to successfully treat either condition. However, although they share significant genetic commonalities, long COVID and ME appear to be best considered as partially overlapping but different diseases.

**Lay Summary:** Myalgic encephalomyelitis (also known as ME/CFS or simply ME), has a debilitating impact on the lives of tens of millions of people around the world. Currently, there are no reliable tests or widely effective treatments for ME. As a result, many doctors don’t recognize or understand the disease, and patients often receive inadequate care. Long COVID has similar challenges. Both ME and long COVID are difficult to study because they cause many different symptoms and affect a lot of different parts of the body. Past genetic research hasn’t found clear answers about why people get ME (or long COVID).

To learn more about what causes ME, we used an approach called the PrecisionLife combinatorial analytics platform, which can find more genetic signals in complex diseases than existing methods. We looked for signatures in people’s genes—specifically, sets of 1 to 4 small changes in DNA (called SNPs)—that together are more common in people who have ME than in healthy people. Such genetic factors may influence how a patient responds to disease triggers such as viral infections, including whether they go on to develop post-viral disease as well as its symptoms and severity.

We studied data from DecodeME participants (cases) and compared them to people who do not have ME (controls) from UK Biobank. We showed that DecodeME participants occur much more often in the group of people with the most genetic signatures compared to groups of people with fewer signatures. We also looked for similarities between the genes we found to be linked to ME and those we had previously linked to long COVID. Finally, we searched the new ME genes we’d found to see which already had drugs that target them in other diseases. We wanted to identify existing medicines that are likely to be effective for some ME patients and show that a simple genetic test might be useful in finding the patients who’ll benefit from that drug.

We found over 22,000 genetic signatures that are linked to a higher risk of ME. The more of these signatures someone has, the more likely they are to have the disease. People with the highest number of signatures had a 1.64 times greater chance of having ME than those with the lowest number. These signatures are connected to over 2,300 different genes, showing that ME involves many genes working together. Out of those, 259 candidate core genes seem to play the largest roles, affecting things like the immune system, how brain and nerve activity is regulated, inflammation, and how the body’s cells respond to damage and pass signals to regulate their processes via the calcium channels. These disease mechanisms are important as they are what we aim to affect with drugs, and all of these mechanisms have had drugs successfully developed for them previously.

This research gives us a much clearer picture of the genetic features that may lie behind ME and could help develop better ways to diagnose and treat it. The complexity of the disease and the fact that patients have different mechanisms driving their disease means though that it is very unlikely that one drug will work for everybody. Understanding which disease mechanisms are involved for each individual patient could make treatments more personalized and help future medical trials to identify patients who will respond more positively to a specific drug.

76 of the 180 genes known to be linked to long COVID are also linked to ME. Some of these genes could lead to new treatments. Since ME and long COVID share many genetic features, there may be some treatments that work for both diseases, but our results indicate they should still be considered as different conditions.

This means that it may be possible to develop treatments that would help some subgroups of both ME and long COVID patients, but it is also likely that some treatments would be specifically useful for just ME cohorts rather than long COVID (and vice versa). These may be completely new drugs or reusing (repurposing) existing drugs for specifically targeted groups of patients based on mechanism, which can be quicker.

These findings are just the beginning, but they show how useful big studies like DecodeME can be and the foundational results that they can enable. We hope these will encourage more research and support from, and for, patients.

## Introduction

### Myalgic Encephalomyelitis and Long COVID Background

Myalgic encephalomyelitis (also known as ME/CFS or simply ME) is a complex, chronic disease characterized by post-exertional malaise (PEM, sometimes referred to as post-exertional neuroimmune exhaustion PENE (Carruthers *et al*. 2011) –– in which symptoms disproportionately worsen, or arise, following minimal physical or mental exertion relative to pre-sickness), as well as neurological components (e.g., unrefreshing sleep, pain, neurocognitive impairment, sensory disturbance), evidence of and cognitive impairment immune/gastro-intestinal and/or genitourinary impairment, and of impairments to energy metabolism/ion transport. Patients may experience a wide spectrum of other symptoms and comorbidities affecting multiple body systems, including dysautonomia, orthostatic intolerance and postural tachycardia, fibromyalgia, IBS, clinical depression, mast cell disease, and connective tissue differences.

ME has a pronounced sex bias, affecting females 3-4 times more frequently than males. Onset of the condition is often linked to an infection (e.g., EBV, enteroviruses, SARS-COV) or other trigger (Chu *et al*. 2019). Many people have gone on to meet the ME diagnostic criteria after developing long COVID (Davis et al., 2023). Approximately 25% of ME patients have symptoms so severe that they are left house- or bed-bound (25% ME Group 2025).

Despite its debilitating impact on the lives of 25-70 million patients worldwide (Vardaman and Gilmour 2025) and its huge societal and economic cost, estimated at over £20B in the UK (Action For ME, in press) and at least $18B - $51B in the US (Centers for Disease Control and Prevention 2025), both scientific research and effective care pathways for ME have been notably lacking (Mirin, Dimmock and Jason 2020; Pheby *et al*. 2020; Thoma *et al*. 2023). This means that the underlying biological mechanisms responsible for ME are still not well understood, posing significant challenges in improving diagnosis and finding effective treatments for patients.

Long COVID is a similarly heterogeneous condition that is estimated to have affected 400 million people worldwide and to cost $1 trillion – 1% of global GDP – annually in healthcare costs and lost productivity (Al-Aly *et al*. 2024a). The World Health Organization (2022) defines long COVID as the continuation or development of new symptoms three months after SARS-CoV-2 infection, that last for at least two months, with no other explanation. Common long COVID symptoms include fatigue, post-exertional malaise, autonomic dysfunction, cognitive dysfunction and shortness of breath, but over 200 symptoms have been reported that can impact daily life.

### Early Genetic Association Studies

Genetic studies of ME have yielded few reproducible results to date. In part this may be related to the broad diagnostic criteria used for case populations, although there may be other explanations. Early studies of CFS (and not specifically ME) found that risk of developing the condition is increased among people who have family members with CFS, suggesting a possible genetic component (Walsh *et al*. 2001; Underhill and O’Gorman 2006; Albright *et al*. 2011). Estimates of narrow-sense heritability of ME and/or CFS vary greatly however, ranging from 3% to 48% (Sullivan *et al*. 2005; Canela-Xandri, Rawlik and Tenesa 2018; Lakhani *et al*. 2019).

Early genome-wide association studies (GWAS) did not find any replicable genetic loci that were significantly associated with increased or decreased prevalence of ME (Dibble, McGrath and Ponting 2020). This is likely due to the limited statistical power provided by available datasets as well as the multi-factorial basis of ME (Arron *et al*. 2024). Of five ME GWAS studies that used data from UK Biobank (UKB) (Sudlow *et al*. 2015), two found no significant loci (Wei *et al*. 2017; Tanigawa *et al*. 2019), two found one significant variant each (Canela-Xandri, Rawlik and Tenesa 2018; Zhou *et al*. 2018), and one identified a female-specific and male-specific significant variant (http://www.nealelab.is/uk-biobank/).

All five of these studies aimed to identify genetic associations across large numbers of phenotypes in UKB and therefore did not attempt to tailor the case-control criteria specifically for ME. None of the four reported variants, two of which are very rare (minor allele frequency < 0.5%) and another of which is multi-allelic and may represent a genotyping artefact, were able to be replicated across multiple studies even though they all relied on the same base dataset (Dibble, McGrath and Ponting 2020).

Two additional GWAS analyses that relied on extremely small and underpowered datasets (<50 cases) reported identification of genetic variants associated with ME. In one, none of the reported genetic associations remained significant after applying standard statistical thresholds for GWAS (Smith *et al*. 2011). The second described 15 variants that were reported to have GWAS-significant disease associations (*p* < 5×10^-8^) (Schlauch *et al*. 2016). The latter is very surprising given the very small dataset size and relatively low heritability of ME, and it is unclear if there were appropriate controls in place for confounding signal of relatedness or population substructure, both of which can result in spurious genetic associations (Sul, Martin and Eskin 2018). Again, none of these purported ME disease associations have yet been replicated in other datasets.

### DecodeME

DecodeME is a genetic study that recruited 21,620 UK participants over the age of 16 who reported a clinical diagnosis of ME by a healthcare professional (Devereux-Cooke *et al*. 2022). DecodeME applied rigorous screening criteria to participants’ questionnaire data to identify participants who pass international consensus criteria for ME and who lack any alternative diagnoses that might be responsible for post-exertional malaise, a unique diagnostic feature of ME (DecodeME 2025). Participants provided self-collected saliva samples that were genotyped using the UKB Axiom array.

DecodeME’s recent pre-print of an initial GWAS comparing 15,579 DecodeME cases with European ancestry and 259,909 UKB controls reported 8 loci significantly associated with ME in that cohort (DecodeME 2025). Annotation of these loci implicated both immunological and neurological processes in ME disease biology.

None of these loci were significantly associated with ME-like traits in a replication study of 15,251 cases and 1,878,066 controls assembled across seven independent biobanks. Four of the loci were associated (*p* < 0.05) with post-exertional malaise and fatigue in UKB and the Netherlands Lifelines biobank, although these associations were not statistically significant after false discovery rate (FDR) corrections for multiple testing. The authors of the DecodeME GWAS posit that this lack of replication may reflect the looser case definition criteria used in those replication data sources relative to DecodeME.

### Combinatorial Analytics Studies of ME

The PrecisionLife® combinatorial analytics platform uses a hypothesis-free approach to identify combinations of features, termed ‘disease signatures’, that are significantly enriched in cases relative to controls or vice-versa. Unlike GWAS, which assumes that each genetic variant acts independently of all the others, combinatorial analysis captures linear and non-linear interactions between combinations of features such as SNP-genotypes and potentially other exogenous factors (Gardner 2021). As such this approach can identify many more significant genetic associations and related biological mechanisms, even in smaller datasets than those required by GWAS (Das *et al*. 2021, 2022b; Taylor *et al*. 2023; Sardell *et al*. 2025a).

The pipeline employed for combinatorial analysis is described in detail elsewhere (Das *et al*. 2021, 2022b). Briefly, it employs a deterministic heuristic algorithm and a hypothesis-free data analytics framework encompassing integrated network geometry and statistical genetics approaches. These are employed over multiple rounds of increasing combinatorial complexity.

During the signature mining phase, the algorithm iteratively combines increasing numbers of features, resulting in multi-SNP disease signatures where each component SNP-genotype contributes to higher disease association. The platform validates these signatures by comparing their properties (e.g., case prevalence, odds ratios) and the properties of their corresponding SNP networks (i.e., the set of all signatures containing a shared SNP – termed the ‘critical’ SNP in the context of that network) against the properties of signatures and SNP networks identified using the same combinatorial analysis methodology across 1,000 fully random permutations of the case-control sample labels.

Statistically validating disease associations for combinatorial disease signatures often requires much larger datasets than replication of GWAS results due to the low frequency of samples having the specific combinations of multiple SNP-genotypes. However, as well as direct biological validation of many targets, the disease signatures, novel candidate genes, and potential drug repurposing candidates that were identified by the PrecisionLife combinatorial analysis approach for complex, heterogenous diseases such as long COVID and endometriosis have been shown to be significantly more likely to also be linked to disease across independent multi-ancestry datasets than corresponding GWAS results (Sardell *et al*. 2025c, 2025a).

A previous combinatorial analysis of ME in a UKB cohort identified 84 disease signatures comprised of 199 SNPs and 14 genes that were significantly enriched in ME patients relative to controls (Das *et al*. 2022b). That study further used those disease signatures to stratify patients into 15 non-exclusive clusters representing potential mechanistic subtypes of ME. These subtypes centered around genes linked to cellular mechanisms hypothesized to underpin ME, including infection response, autoimmune development, mitochondrial dysfunction, and neurotransmitter biology, and circadian rhythm.

These 84 signatures were then used in a study of actively protective biology that identified 276 ME protective signatures and 9 disease resilience genes that were significantly enriched in UKB controls who remain healthy despite possessing many ME disease risk signatures – dubbed the ‘high-risk protected’ cohort (Sardell, Das, Taylor, et al., 2025).

A combinatorial analysis of long COVID patients from the Sano GOLD cohort identified overlap between the mechanisms associated with Severe and Fatigue Dominant long COVID phenotypes (73 genes found) and the mechanisms linked to ME in UKB, including 9 shared genes (Das *et al*. 2022b). The results of the long COVID combinatorial analysis were recently reproduced with 92% overlap of genes by the authors of the original study in a cohort of mixed ancestry long COVID patients from the All of Us dataset (Sardell *et al*. 2025c).

An independent study likewise found that 12 out of 80 of the ‘critical’ SNPs and 4 genes identified by the long COVID combinatorial analysis from Das et al. (2022b) were also associated with long COVID in at least one of four patient datasets (Cheng 2025). This rate of replication is likely underestimated as Cheng (2025) evaluated each SNP individually rather than the networks of disease signatures containing each SNP, which better reflect complex non-linear disease biology.

### Aims of Study

This study formed part of the Innovate UK funded LOCOME project (#10083274), investigating both long COVID and ME. The intention of the project was to compare the use of combinatorial analysis alongside GWAS to improve understanding of the diseases’ biology, inform new diagnostic tests, and identify potential drug repurposing candidates.

We applied combinatorial analytics to populations from the DecodeME dataset to identify disease signatures and genes associated with increased risk of ME. The larger sample numbers and rigorous phenotypic screening employed in the construction of the DecodeME patient cohort allowed us to identify many more genetic associations relative to the previous combinatorial analysis, which relied solely on UKB patient data with its less consistent diagnostic criteria from non-disease specific participant surveys.

The combinatorial analytics approach allowed identification of many multiply reproducible genetic associations not found by GWAS. These findings were further refined by assessing the reproducibility of the disease signatures and their component SNPs in a UKB cohort as well as in independent case cohorts of participants from DecodeME. This resulted in a set of multiply reproducible signatures that can potentially be used to inform precision medicine patient stratification tools and test new drug repurposing candidates for ME. We also recapitulated and identified broader overlap between genes implicated in ME and those previously associated with long COVID.

## Materials and Methods

### Study Datasets

DecodeME provided genotype data for 14,767 participants who satisfied the study’s inclusion criteria based on the Canadian Consensus and/or US Institute of Medicine / National Academy of Medicine criteria for ME/CFS using survey responses, who had received a diagnosis of ME, CFS, ME/CFS or CFS/ME from a health professional, and who consented to data sharing with accredited research teams. DecodeME participants were genotyped in four batches (DecodeME 2025), but only the first three were included in this analysis due to the timing of data availability.

Although all were genotyped using the Axiom UK Biobank array, non-trivial variations in quality control (QC) resulted in major differences in SNP counts between the DecodeME batches, and a substantial reduction in the number of participants (cases) and SNPs that could be used in the study. The extensive QC processes that were undertaken to remove batch and population structure effects and other data quality issues are described in the Supplementary Methods section.

In these batches between 82-86% of DecodeME cases and 53-59% of the DecodeME SNPs were able to be used for this analysis. Total counts of cases and SNPs for each batch used in this study are listed in Table 1 below. The number of females and males in the cohorts are listed in Supplementary Table 2.

**Table 1.** Number of cases and SNPs from the DecodeME and UKB cohorts before and after LOCOME Study QC, broken down by batch.

| ME Cohort | # Cases<br>before QC | # SNPs<br>before QC | # Cases<br>after Study QC (%) | # SNPs<br>after Study QC (%) |
| --- | --- | --- | --- | --- |
| DecodeME Batch 1 | 3,944 | 772,374 | 3,405 (86%) | 408,873 (53%) |
| DecodeME Batch 2 | 4,349 | 656,807 | 3,677 (85%) | 362,798 (55%) |
| DecodeME Batch 3 | 4,238 | 645,542 | 3,487 (82%) | 381,756 (59%) |
| DecodeME Batch 2+3 Split 1 |  | -- | 3,585 | 349,734 |
| DecodeME Batch 2+3 Split 2 (Test) |  | -- | 3,579 | 350,014 |
| UK Biobank –<br>Pain Questionnaire | 2,382 | 554,063 | 1,985 (83%) | 519,337 (94%) |

### UK Biobank Controls

The DecodeME dataset is restricted to participants who satisfy the clinical case criteria for ME, CFS, ME/CFS or CFS/ME (Devereux-Cooke *et al*. 2022; DecodeME 2025). We therefore relied on UKB as a source of controls for the genetic association analyses.

We used the same criteria as Das et al. (2022b) to identify potential UKB controls, i.e., participants with no evidence in UKB’s Hospital Episodes Statistics (HES), primary care, or self-reported data fields indicating diagnoses of chronic fatigue, post-exertional malaise, post-viral fatigue syndrome or myalgia (Supplementary Table 1). We further excluded all UKB participants previously used as controls in the combinatorial analyses described in Das et al. (2022b). Ensuring we only used non-overlapping controls allowed us to use the previously identified UKB ME dataset for independently assessing the reproducibility of the output of the DecodeME combinatorial analysis.

### Creation of Merged Datasets

Our analytical pipeline used four cohorts of ME cases and controls (Figure 1):

1. A DecodeME ‘Discovery’ dataset used to detect disease signatures via hypothesis-free combinatorial analysis of a cohort of DecodeME cases and UKB controls.
2. A ‘UKB Refinement’ dataset used to filter disease signatures and component SNPs based on reproducibility of disease associations in an independent cohort of UKB cases and controls.
3. A ‘DecodeME Refinement’ dataset used to filter disease signatures and component SNPs based on reproducibility of disease associations in a cohort of independent DecodeME cases and the same controls as the UKB Refinement dataset.
4. A ‘Test’ dataset used to evaluate the predictivity of the final set of ‘double-refined’ signatures (from the UKB Refinement and DecodeME Refinement steps respectively) in an independent cohort of DecodeME cases and UKB controls not included in any other dataset.

**Figure 1.**
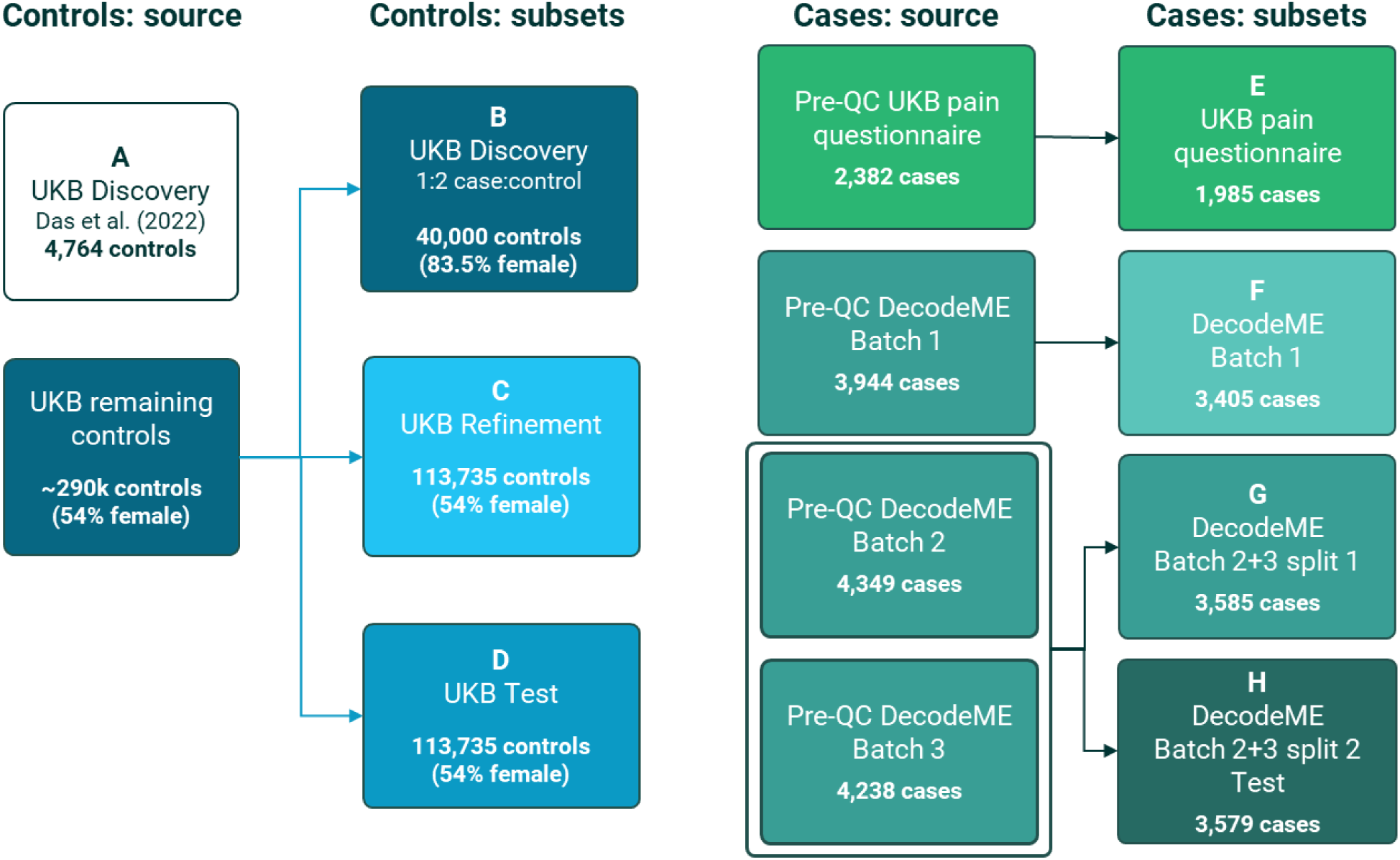
Study cohorts used as sources of ME cases and controls in the disease signature Discovery, Refinement, and Testing pipeline and the cohort datasets used for Analysis 1 and Analysis 2

#### Identification of UKB control cohorts and UKB refinement cases

We began by identifying non-overlapping sets of UKB controls to use for Discovery, Refinement, and Testing.

Control Cohort B: We first set aside 40,000 samples from the set of potential UKB controls to use for disease signature Discovery (i.e., combinatorial analyses). This number was selected to potentially allow for a minimum 1:2 case:control ratio in a combinatorial analysis of the full DecodeME cohort prior to knowing how many participants had consented to data sharing. The set of Discovery controls was selected to be 83.5% female to match available data on the sex-ratio in the DecodeME cohort.

Control Cohort C: To create a set of Refinement controls, we first randomly selected half the remaining female UKB potential controls (i.e., excluding Discovery controls). We then randomly selected remaining male UKB potential controls to create a cohort of UKB Refinement controls that is 54% female, matching the overall female:male ratio in UKB. For the UKB Refinement dataset (Step 2), we paired the UKB Refinement controls with ME cases identified using UKB’s pain questionnaire (as described in Das *et al*. 2022b).

Control Cohort D: Finally, we created a set of UKB Test controls by taking the remaining potential female UKB potential controls and merging them with randomly selected remaining male UKB potential controls to again produce a controls cohort that is 54% female.

#### Creation of DecodeME Discovery, Refinement, and Test datasets

We conducted two complementary analyses to fully utilize the available samples from DecodeME (Figures 1-2), relying on three independent cohorts of DecodeME participants.

**Figure 2.**
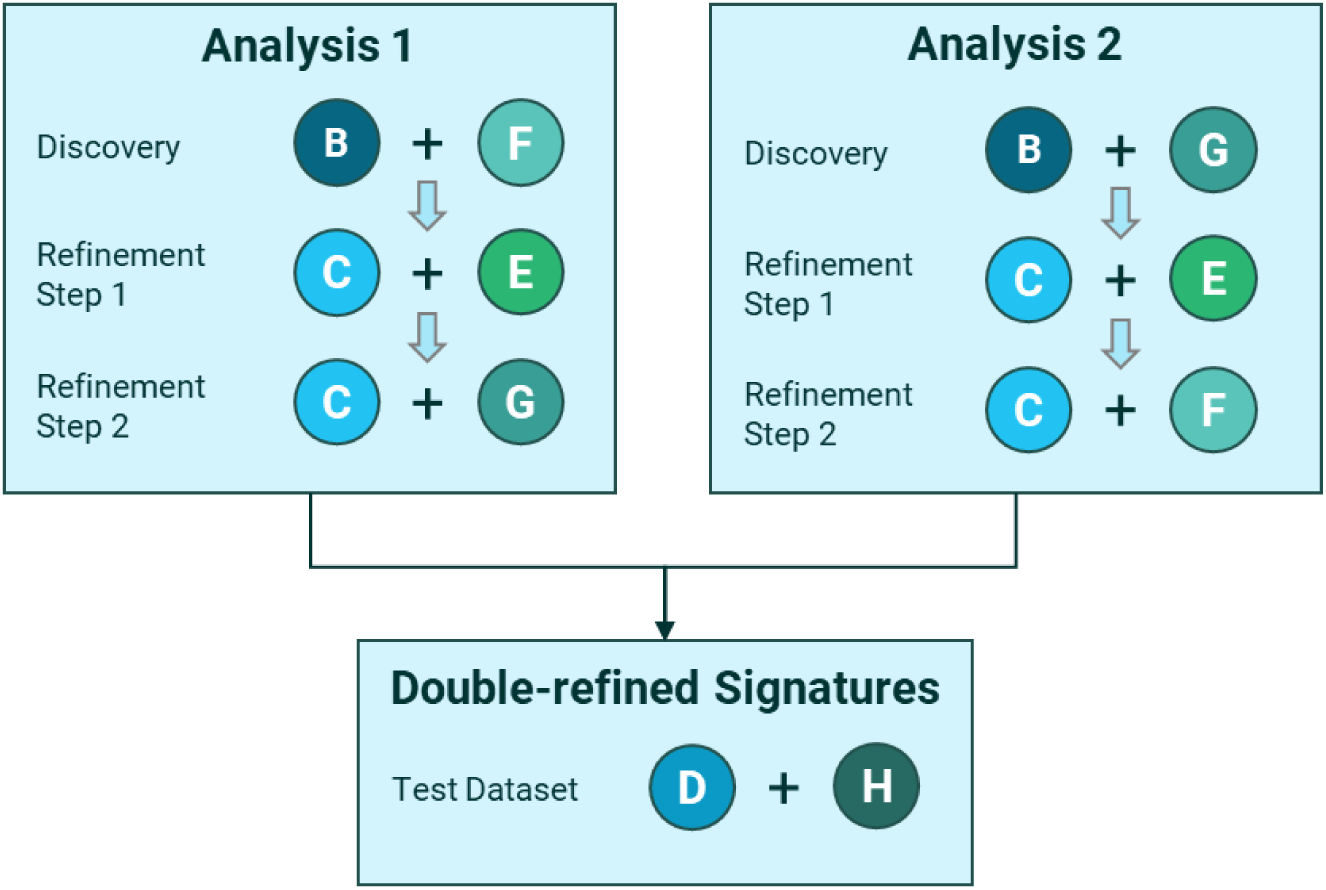
Disease signature Discovery, Refinement, and Testing pipelines and cohort datasets used for Analysis 1 and Analysis 2

Case Cohort F was comprised of ME participants from DecodeME Batch 1 (which was the first cohort available for analysis). Case Cohorts G and H were comprised of ME participants from DecodeME Batches 2 and 3 (which were made available after the initial combinatorial analysis had been performed). To mitigate any potential batch effects between Batches 2 and 3, we merged their QCed cases. We then re-split the cases in half to form Case Cohorts G and H, ensuring a consistent ratio of cases from Batch 2 vs. Batch 3 and females vs. males within each batch and case cohort. Case Cohort G was used for Discovery in Analysis 2 and Refinement in Analysis 1, while Case Cohort H was merged with the UKB Test controls (Cohort D).

#### Combinatorial Analyses

In Analysis 1, we performed combinatorial analysis using a Discovery dataset comprised of the UKB Discovery controls (Controls B) paired with ME cases from DecodeME Batch 1 (Cases F) (see Figure 2). Refinement was first conducted in the UKB Refinement dataset (Controls C paired with Cases E) and then in a Refinement dataset comprised of the UKB Refinement controls (Controls C) paired with the first DecodeME Batch 2+3 split (Cases G).

In Analysis 2, we inverted the study design from Analysis 1. First, we performed combinatorial analysis using a Discovery dataset comprised of the UKB Discovery controls (Controls B) paired with the first DecodeME Batch 2+3 split (Cases G). Refinement was first conducted in the UKB Refinement dataset and then in a Refinement dataset comprised of the UKB Refinement controls (Controls C) paired with DecodeME Batch 1 cases (Cases F).

Pooling the output of Analysis 1 and Analysis 2 provides a set of ‘double-refined’ disease signatures that are associated with ME across three cohorts (i.e., a DecodeME Discovery dataset, a UKB Refinement dataset, and a DecodeME Refinement dataset).

Note that after QC, many SNPs that were present in the Analysis 1 Discovery dataset derived from DecodeME Batch 1 were missing from the Test Dataset and Analysis 1 Refinement datasets derived from DecodeME Batches 2+3. We therefore imputed these missing SNPs using the approach described in the Supplementary Methods to allow for refinement and validation of signatures containing them.

### Dataset QC

We conducted standard quality control (QC) individually on each batch of participants provided by DecodeME and each merged dataset based on the protocol described in Marees et al. (2018). Details are provided in the Supplementary Methods.

Population substructure within a cohort can result in false positive gene-disease associations when disease prevalence is correlated with patient ancestry, for example if ME is underdiagnosed in non-European genetic ancestry patients. In this scenario, genotypes that differ in frequency between historical human populations either due to selection or genetic drift can be indirectly associated with disease even though they do not directly affect disease biology.

Due to the very high proportion of European ancestry participants and low representation of other ancestries, we filtered the DecodeME cohorts to include only participants classified as having European ancestry (see Supplementary Methods). This reduced the potential that the disease signatures associated with increased prevalence of ME in DecodeME are artefacts of population substructure rather than reflecting genetic features directly relevant to disease biology.

Merging DecodeME cases with UKB controls introduces the potential for batch effects to produce large numbers of false positive disease associations (Zuvich *et al*. 2011; Sinnott and Kraft 2012; Tom *et al*. 2017). That is, differences in signature frequency between cases and controls may reflect technological artefacts in the data rather than differences in disease biology.

Although DecodeME used the same genotyping array as UKB, other differences in the genotyping pipeline (e.g., in sample collection, sample preparation and storage, DNA extraction, machine calibration, genotype calling, or bioinformatic pipelines) could result in material differences in genotyping error rates and observed allele frequencies between the two datasets.

Because case-control status is perfectly correlated with the dataset source in our data, identifying and removing these batch effects while retaining true genetic associations is challenging. We therefore undertook a series of extensive and highly conservative QC analyses to identify and remove false positive signal associated with differences in allele frequency between DecodeME and UKB and between DecodeME batches that reflect genotyping artefacts. These QC processes are described in detail in the Supplementary Methods.

### Analytical Pipeline – Signature Discovery and Refinement

We applied a multi-step analytical pipeline to identify and refine disease signatures for ME using non-overlapping cohorts of ME cases and healthy controls (see Figure 3). We began by using the PrecisionLife combinatorial analytics platform to identify disease signatures significantly enriched in ME cases relative to UKB controls in one of the two Discovery datasets.

**Figure 3.**
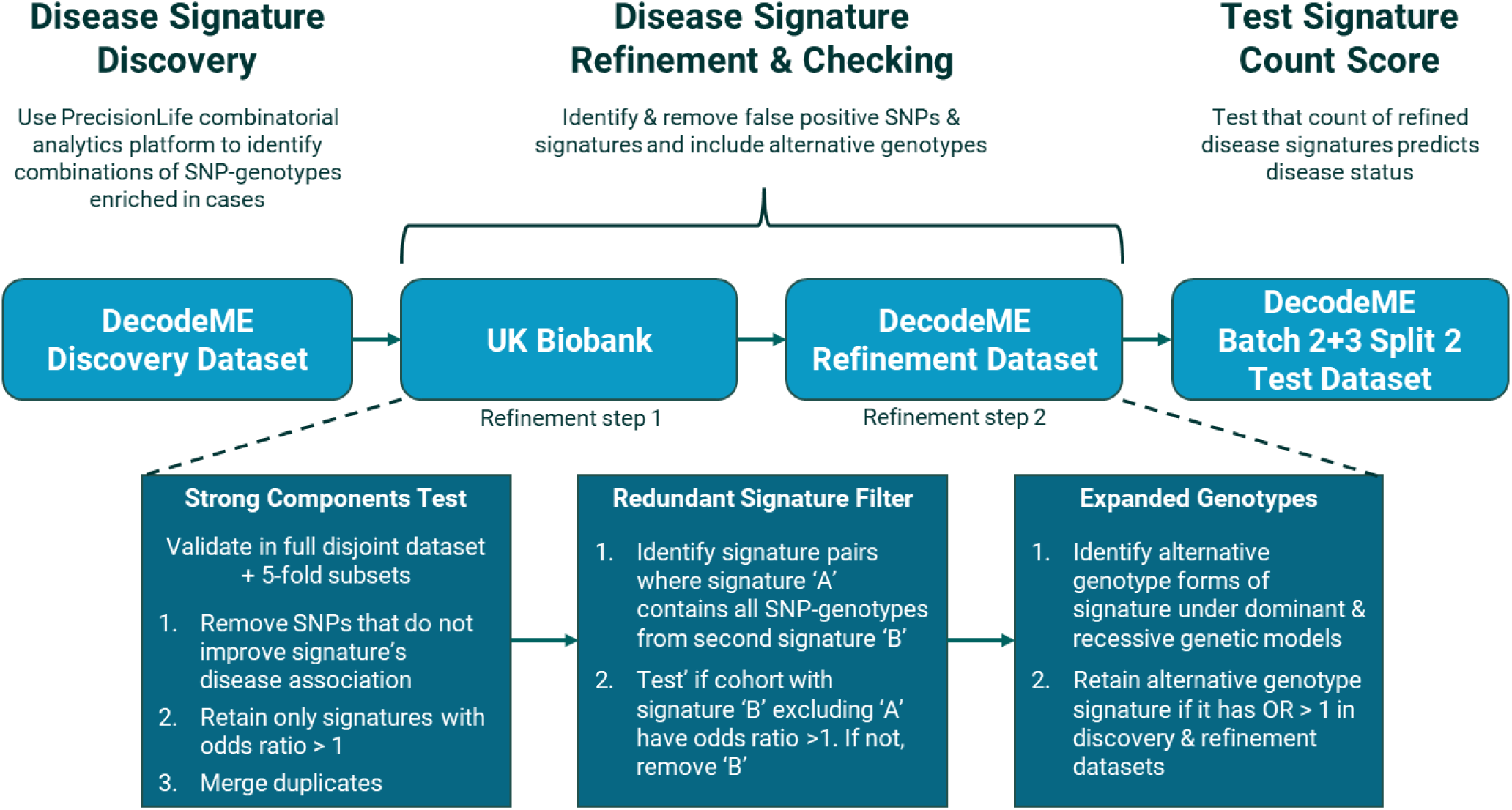
Analytical pipeline for Discovery and Refinement of disease signatures and Testing of signature count score for ME. The Discovery and Refinement datasets are illustrated in Figure 1. These processes are described further in the Supplementary Methods section.

After combinatorial analysis, we applied a process we call ‘Refinement’ to the set of disease signatures (see Figure 3 and Supplementary Methods section for more detail). This entailed:

1. Removing signatures and/or component SNP-genotypes that do not show reproducible associations with ME in a second cohort (‘strong components test’).
2. Removing signatures that do not provide insight into disease associations that are independent of other signatures (‘redundant signature filter’).
3. Expanding the set of genotypes reflected in the disease signatures to reflect standard genetic models when appropriate (‘expanded genotypes’).

### Analytical Pipeline – Testing of Predictive Power

The above pipeline resulted in a set of ‘double-refined’ disease signatures that were associated with increased odds of ME across three independent cohorts of participants (one Discovery + two Refinement cohorts). To evaluate the aggregate predictive power of these signatures, we tested whether the count of double-refined signatures possessed by a patient is correlated with prevalence of ME in the Test dataset, which was comprised of cases and controls not previously used for Discovery or Refinement in either analysis. For each patient, we calculated a signature count score reflecting a simple count of the number of signatures possessed by each patient.

As noted in the Results section, we identified thousands of correlated disease signatures containing at least one SNP located within the *6p22.1* locus, which strongly influenced the signature count. We therefore excluded these signatures from the set of signatures evaluated by the signature count score.

We then calculated a logistic regression using the *glm* function in R with CRS as the independent variable, case-control status as the dependent variable, and genetic sex and 10 genetic principal components (PCs) as confounders to evaluate whether the signature count score is significantly associated with case-control status (see Supplementary Methods).

We further sorted individuals in the Test dataset by signature count score and calculated the odds of ME (i.e., # cases / # controls) for each signature count score decile. Odds ratios were calculated by dividing the odds for each decile by the odds of ME for individuals falling within the bottom 10% of signature count score values.

### Gene Annotation and Prioritization

SNPs identified in the disease signatures were conservatively mapped to protein-coding genes based on the human reference genome (GRCh37). Only SNPs located within the transcription start and stop positions of genes were mapped to the corresponding gene(s). These genes were then used for overlap analyses with other studies.

To aid in identifying the candidate core, potentially causal, genes and mechanisms of action (MoAs) linked to ME, we used the appropriate DecodeME Refinement datasets (C+F and C+G) to filter the mapped genes based on the strength of their associations with ME and the prevalence of associated disease signatures in ME patients. We then constructed networks for each SNP comprised of the set of all double-refined disease signatures that contained that SNP (i.e., the network’s defining SNP). Cases and controls were assigned to a network if they possessed at least one of the constituent disease signatures. This allowed us to calculate a network odds ratio reflecting the odds of ME for samples within and not within each network, and each network’s case prevalence (i.e., the fraction of total ME cases who were assigned to the network).

We further calculated a SNP odds ratio reflecting the odds of ME for a network relative to the odds of ME for a matching network comprised of the same constituent signatures but with the defining SNP removed, along with a corresponding Fisher’s exact test *p*-value. This measure allows us to quantify the association with ME for individual SNPs in a combinatorial setting.

Finally, we identified the set of SNPs that define networks with SNP odds ratio *p-*value less than 0.05 and a case prevalence greater than 20%. Genes mapped to these SNPs, and those with a minimum SNP network odds ratio exceeding the median value across all genes were identified as the candidate core gene set. Supplementary Table 3 shows the varying SNP and candidate core gene counts at different prevalence thresholds.

All genes were annotated using data from over 50 public data sources (see Supplementary Table 4) to characterize their biological roles and potential mechanism of action link to ME. We first performed pathway enrichment analysis on the candidate core genes using Gene Set Enrichment Analysis (GSEA) (Subramanian *et al*. 2005) with Gene Ontology Biological Process (GO-BP) (Aleksander *et al*. 2023) and WikiPathways annotations (Agrawal *et al*. 2024).

### Overlap with Long COVID

We tested for overlap between the genes identified in our hypothesis-free ME analyses and the genetic associations for long COVID that were previously identified by combinatorial analysis in the Sano GOLD cohort and which also exhibited high rates of reproducibility (92%) in an All of Us cohort (Taylor *et al*. 2023; Sardell *et al*. 2025c). These hypothesis-free studies identified genes associated with long COVID in three partially overlapping cohorts from the Sano GOLD dataset (Taylor *et al*. 2023):

1. Patients who have a ‘Fatigue Dominant’ form of long COVID
2. Patients who have a ‘Severe’ form of long COVID
3. Patients who satisfy the ‘General’ criteria for long COVID

We used a Fisher’s exact test to determine whether we observed a significant enrichment of long COVID genes that also map to the double refined ME disease signatures.

We also tested this overlap using an independent method – i.e. not relying directly on the PrecisionLife combinatorial analytics methodology which may be biased towards outputting larger genes that map to many SNPs on the Axiom UKB genotyping array. To do this, we identified the overlap between the long COVID genes reported in the hypothesis-free analyses above and the output of an independent machine learning (ML) analysis that quantified the relative feature importance of SNPs and genes in models that classify people into ME cases and controls.

The accuracy and robustness of the ML model for ME is limited due to the large numbers of included SNPs and their relatively small feature importances. It would not be useful as a discovery tool in its own right, but it does allow identification of genes previously associated with long COVID that are also highly associated with ME.

We performed two analyses using the ML model for ME. First, we ranked SNPs by mean feature weight in the ML model for ME and tested whether SNPs mapping to the long COVID genes had significantly stronger ranks (i.e., high feature importances) relative to comparable sets of random genes. To test for potential biases in the methodology, we contrasted the results of the overlap analysis for long COVID genes with the same analysis for genes identified by combinatorial analyses of endometriosis (Sardell *et al*. 2025a) and coronary artery disease (CAD) (<u>unpublished results</u>, Supplementary Table 5). Second, we identified genes mapping to a small set of SNPs with very high feature importances in at least one of the ten folds of the ML model for ME, then tested whether we observed significantly enriched overlap between these and the long COVID genes.

Details of these analyses and the ML model are included in the Supplementary Methods.

### Patient and Public Involvement and Engagement (PPIE)

A PPIE advisory panel of individuals with lived experience of ME and/or long COVID and patient advocates was established in collaboration with the charity Action for ME. Members were recruited through the charity’s networks and reimbursed according to NIHR payment guidelines.

The panel met virtually 25 times during the LOCOME project and contributed particularly to both the planning and operational aspects of the study reviewing progress on a monthly basis ensuring clarity in planning future work. In addition, they brought unique perspectives and critical thinking from lived experience and extensive knowledge from the wider patient and academic community, improving analysis and written communications of the results.

## Results

### Combinatorial Analysis of the DecodeME Cohorts

We identified 22,411 double-refined signatures, comprised of 7,555 SNPs mapped to 2,311 genes, that are consistently associated with increased odds of ME in multiple DecodeME and UKB cohorts (Table 2, Supplementary Tables 6-7). Ignoring genotypes, the double-refined signatures represent 20,496 combinations of SNPs. 38% of the double-refined signatures have odds ratios greater than 1.1 in the appropriate Refinement dataset, with a maximum odds ratio = 2.55, mean odds ratio = 1.10, and median odds ratios = 1.09 (see Figure 4, Supplementary Figure 4).

**Figure 4.**
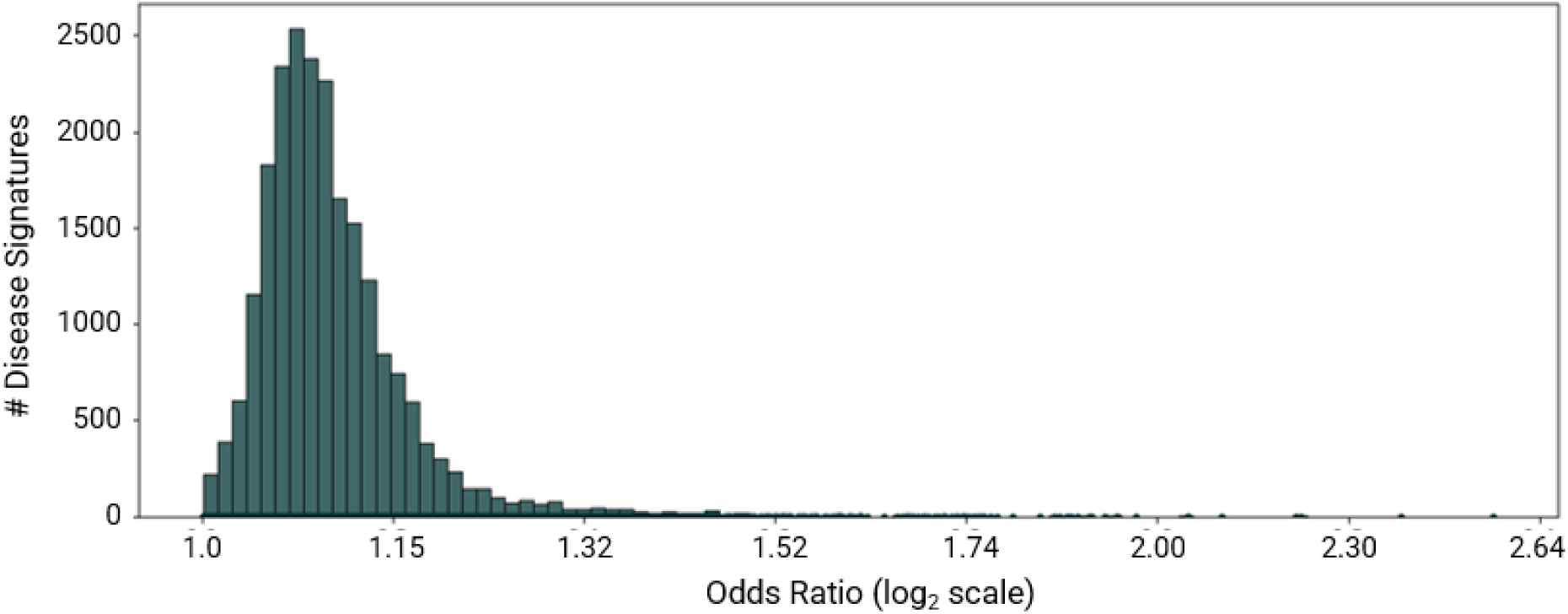
Distribution of odds ratios of double-refined signatures in appropriate Refinement datasets.

**Table 2.** Number of signatures, component SNPs, and mapping genes from the initial DecodeME combinatorial analyses after Refinement in UKB and an independent DecodeME cohort. Candidate core genes were selected based on association with ME and case prevalence in a DecodeME Refinement dataset.

| # Double-refined signatures | # SNPs in double-refined signatures | # Genes mapping to double-refined signatures | # Candidate core genes |
| --- | --- | --- | --- |
| 22,411 | 7,555 | 2,311 | 259 |

Extended Table 1 lists the 7,555 SNPs and 2311 genes mapped to the double-refined signatures and Extended Table 2 lists locus information for the SNPs and signatures. The SNP combinations predominantly have a ratio of females to males that is closely aligned to the overall sex ratio in the dataset, suggesting that the signatures are not sex-biased (Supplementary Figure 5). Breakdowns of number of signatures and SNPs following each step of the Discovery/Refinement pipeline are included in the Supplementary Materials.

#### Candidate core genes

To aid in identification of candidate core genes and MoAs, we filtered the gene list from each analysis by prioritizing genes that have the strongest disease odds ratio and mapped to at least 20% of cases in the appropriate Refinement dataset (so as to avoid overfitting to the Discovery datasets). In total, 259 unique genes (Extended Table 3) satisfied these criteria.

Pathway enrichment analysis on the candidate core gene set highlighted multiple cellular processes involved in nervous system development and neural signaling, immune response, cellular stress response and calcium signaling (Supplementary Figure 6). Many of these processes have been previously linked to ME, supporting their potential involvement (Das *et al*. 2022b; Annesley *et al*. 2024; Sardell *et al*. 2025b).

This is exemplified by genes such as *TLR3*, involved in innate immune response, which has an agonist currently in clinical development for ME treatment (Strayer, Young and Mitchell 2020), along with *CSE1L and DCC* which were associated with ME and chronic pain respectively in a GWAS including DecodeME participants of European ancestry (DecodeME 2025).

### Aggregate Signal from Disease Signatures

In order to evaluate the predictive power of the double-refined disease signatures, we used the DecodeME Test dataset (cohorts D+H) to test if the count of double-refined signatures is associated with increased odds of ME. Importantly, none of the samples in the Test dataset overlap with samples used for disease signature Discovery or Refinement.

This is not intended to be a polygenic risk score or equivalent – it is a simplistic, non-optimized tool to test whether the cumulative set of signatures are associated with increased risk of ME, while minimizing the risk of overfitting.

Each feature (i.e., signature in this case) is assumed to additively and independently contribute to a patient’s risk of disease, and all signatures are assumed to have identical effect sizes. Combinatorial signatures are clearly not independent, as they can (and often do) share SNPs that are identical or in linkage disequilibrium. Nor do they have identical effect sizes. Nonetheless, this simplistic additive signature count score approach enables unbiased testing of the association of the signatures in aggregate.

We observed a bimodal distribution for the number of double-refined signatures possessed by samples in the Test dataset (Figure 6a). This reflects signatures that contain at least one of 187 SNPs from the extended major histocompatibility (MHC) region located within the *p22.1* cytogenic band of chromosome 6. In total, 6,404 double-refined signatures (29% of total) map to this region of low recombination and are strongly correlated due to shared SNPs and high linkage disequilibrium. As a result, individuals are more likely to possess either many or few signatures mapped to this region than they are to possess intermediate numbers of signatures.

**Figure 6.**
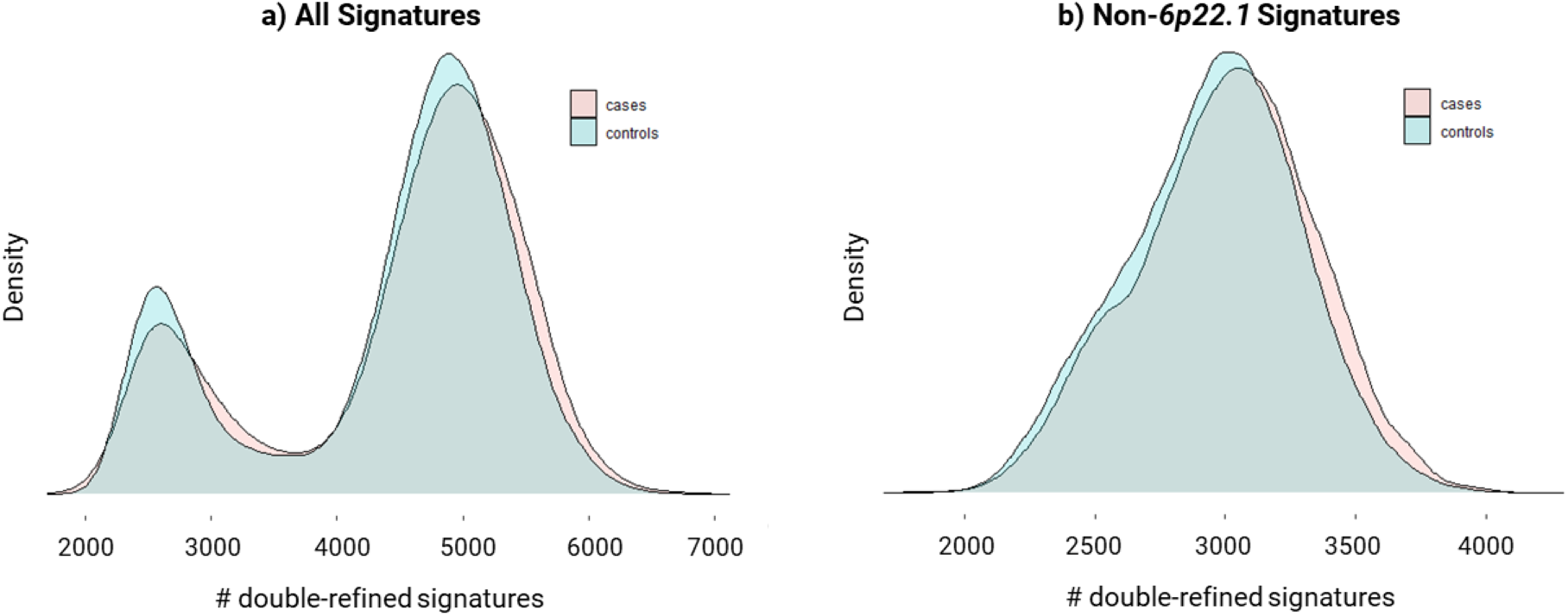
Density plots showing distribution of #number of double-refined signatures possessed by individuals in the Test dataset cohort. Distributions for cases (pink) and controls (blue) are plotted separately. a) Count of all double-refined ME signatures. b) Count of double-refined ME signatures not mapped to *6p22.1*.

Removing signatures that contain at least one *6p22.1* SNP results in a unimodal distribution of double-refined signatures possessed by ME cases (Figure 6b). Below we refer to this adjusted count as the signature count score, which removes the potential strong confounding factor introduced by overweighting signal related to *6p22.1*.

We observed a highly significant correlation between the signature count score and ME in the Test dataset (OR = 1.23 per standard deviation increase, *p* = 4×10^-21^) when including sex and the top 10 genetic principal components as confounders in the logistic regression. The disease odds ratio for individuals with a top 10% signature count score relative to individuals with a bottom 10% signature count score was 1.64 (Figure 7), while the odds ratio for the top 5% vs. bottom 5% was 1.89.

**Figure 7.**
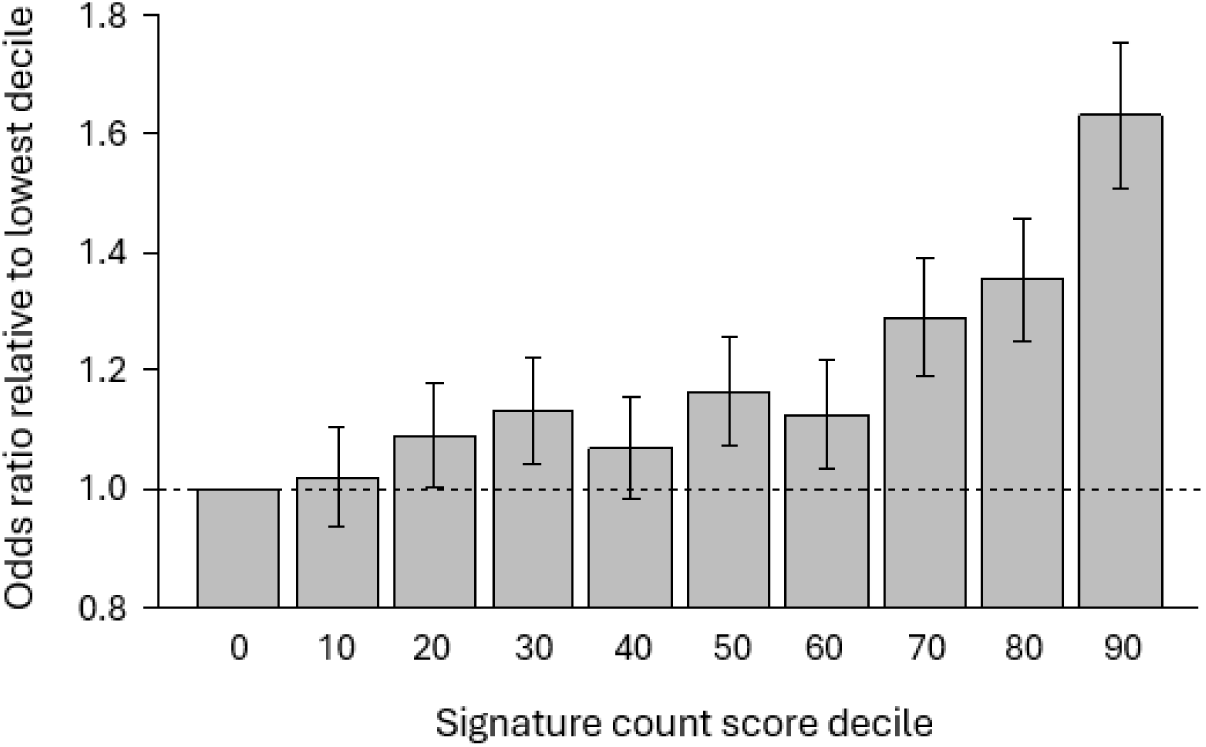
Relative odds of ME across deciles of counts of signatures possessed by individuals in the Test dataset. Odds for each decile are standardized to the odds of the bottom 10% decile. Deciles are labeled with their equivalent lower-bound percentile. Error bars represent standard error. Signature count excludes double-refined ME signatures mapped to *6p22.1* to eliminate confounding due to the disproportionate number of strongly correlated signatures mapped to that locus.

### Overlap with DecodeME GWAS Results

The DecodeME GWAS study identified 8 genome-wide significant loci associated with ME in the DecodeME cohort using 15,579 cases, over 150,000 controls, and more than 8.8 million variants including imputed variants (DecodeME 2025). For each locus, they identified a lead variant mapped to a gene and classified a total of 37 genes into Tier 1 and Tier 2 based on the strength of genetic evidence and biological relevance (DecodeME 2025). The study also reported an additional non-genome-wide significant gene linked to chronic pain.

The double-refined signatures map to 13 of the 32 Tier 1 or 2 DecodeME GWAS study genes including genes located in 6 of the 8 GWAS loci (Table 4, Supplementary Table 8). The double-refined signatures also contain SNPs that map to the remaining 2 loci but are not located within protein-coding genes, potentially representing non-coding variants that affect expression of one or more Tier 1 or 2 genes. Of the Tier 1 or 2 GWAS genes, only *CSE1L* was included among the 259 candidate core genes, along with *DCC,* which is linked to chronic pain.

**Table 4.**
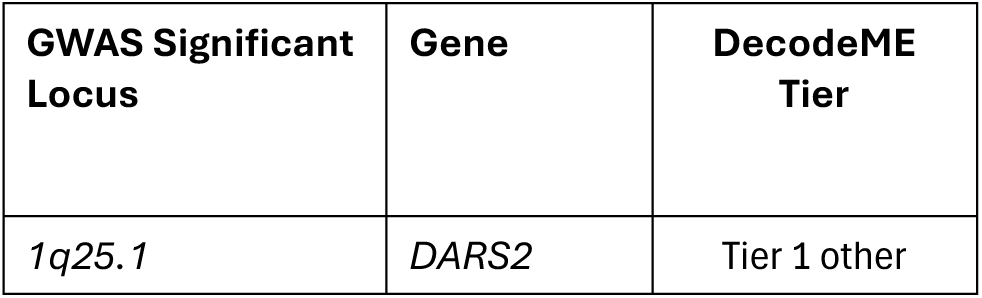
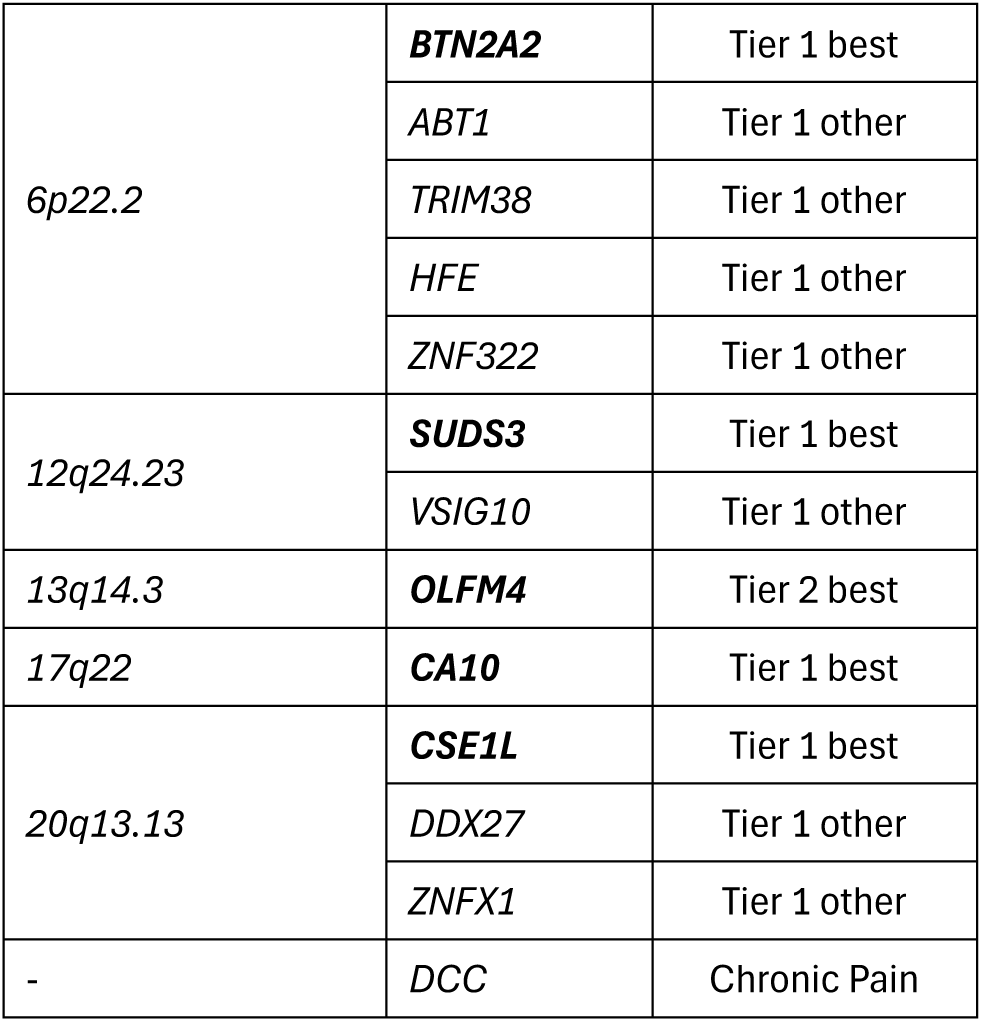
Genes identified by combinatorial analysis that map to GWAS significant and other loci reported by DecodeME study. The lead genes (n=5) reported by the DecodeME GWAS are shown in bold.

| GWAS Significant Locus | Gene | DecodeME Tier |
| --- | --- | --- |
| *1q25.1* | *DARS2* | Tier 1 other |
| 6p22.2 | **BTN2A2** | Tier 1 best |
|  | *ABT1* | Tier 1 other |
|  | *TRIM38* | Tier 1 other |
|  | *HFE* | Tier 1 other |
|  | *ZNF322* | Tier 1 other |
| 12q24.23 | **SUDS3** | Tier 1 best |
|  | *VSIG10* | Tier 1 other |
| 13q14.3 | **OLFM4** | Tier 2 best |
| 17q22 | **CA10** | Tier 1 best |
| 20q13.13 | **CSE1L** | Tier 1 best |
|  | *DDX27* | Tier 1 other |
|  | *ZNFX1* | Tier 1 other |
| - | *DCC* | Chronic Pain |

### Overlap with Long COVID

We previously identified 235 long COVID associated genes in three patient cohorts derived from the Sano GOLD study cohort (Taylor *et al*. 2023), of which 180 were reproduced in All of Us long COVID cohort (Sardell *et al*. 2025c). These included:

1. 43 genes from a ‘Severe’ cohort who reported the most diverse and severe symptoms.
2. 35 genes from a ‘Fatigue Dominant’ cohort who reported mainly fatigue-related symptoms.
3. 165 genes, from a ‘General’ cohort self-reporting ongoing or new symptoms 12 weeks after initial SARS-CoV-2 infection, with symptoms lasting at least 2 months without other explanation.

#### Gene overlap between ME & long COVID disease signatures

102 (43%) of the ME genes previously associated with long COVID also mapped to the double-refined ME signatures, including 76 of the 180 genes (42%) that reproduced in the All of Us long COVID cohort (Supplementary Table 9). Both results represent a significant enrichment of long COVID genes mapping to the ME signatures (Fisher’s exact test *p* < 10^-30^ and *p* < 10^-21^ respectively). These *p*-values, however, assume that all genes have similar likelihood of being observed in a combinatorial analysis. However, genes vary considerably in length, and our gene mapping approach could potentially bias the PrecisionLife platform towards identifying longer genes that map to many SNPs.

We therefore also conducted a second enrichment analysis to evaluate the overlap between ME and long COVID genes, employing a length-matched permutation test to control for potential biases that favor identification of longer genes (see Supplementary Methods). This analysis showed significant enrichment (*p* < 0.016) across all three gene length bins (lengths lower than 50 kb, 50 - 250 kb and above 250 kb) after adjusting for multiple-testing correction (see Supplementary Table 10). This result confirms that the overlap between long COVID and ME genes is not simply an artifact of gene length.

The identified overlap between ME and long COVID is perhaps an underestimate as the originating studies were carried out based on genotype data from two different arrays – Axiom UK Biobank for ME and Illumina Global Screening Array for the Sano GOLD study. The two QCed datasets had very limited SNP overlap (just 10%), and without using imputation, it is only possible to evaluate overlap at a gene level. Since both the DecodeME and All of Us datasets provided fewer than 400,000 SNPs this can only count as a minimum estimate of potential overlap.

#### Machine learning model for ME

We also observed a highly significant enrichment of top ranked SNPs in the ML model for ME (i.e., SNPs with high feature importance) assigned to the sets of genes previously associated with long COVID (Figure 8). The top 25^th^ percentile ranks are significantly higher in the sets of SNPs assigned to the gene sets for each long COVID phenotype than the expected top 25^th^ percentile rank for randomly selected sets of the same numbers of genes (*p <* 0.001 for all three long COVID phenotypes).

**Figure 8.**
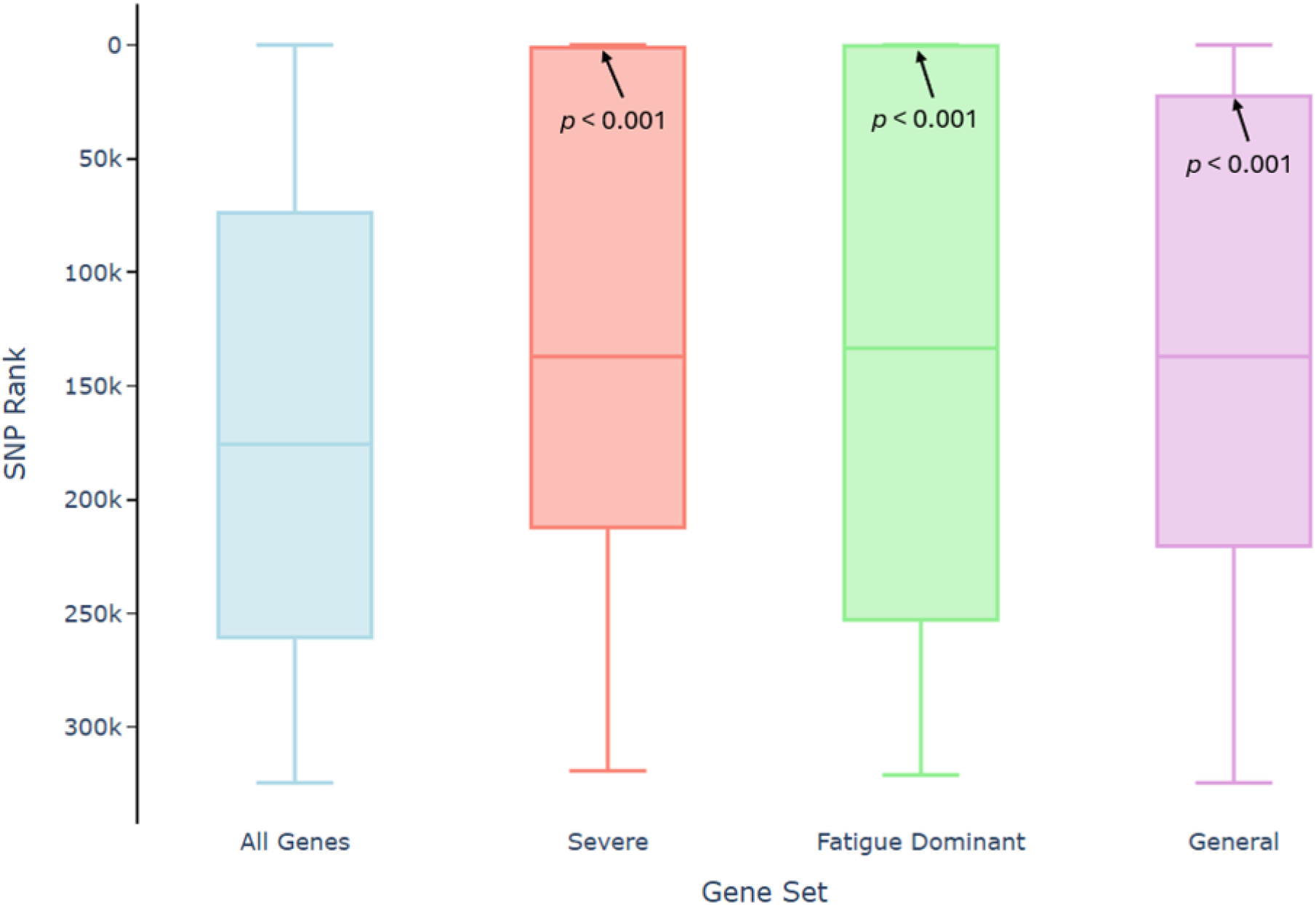
SNP ranks in the ML model for ME for all SNPs assigned to autosomal protein-coding genes (left) and SNPs assigned to genes associated with Severe, Fatigue Dominant, and General long COVID phenotypes in the Sano GOLD cohort (right three boxplots). Higher SNP ranks (i.e. higher positions on the y-axis) denote high feature importance in the ML model. Boxes represent the top 25^th^ and bottom 25^th^ percentile ranks for SNPs assigned to each gene set with solid line denoting median SNP rank. *p*-values are the probability of randomly observing a top 25^th^ percentile SNP rank that high based on 10,000 gene subsets of equivalent size randomly sampled from the full list of autosomal protein-coding genes.

SNP rankings in the ML model were particularly strong for the genes previously associated with Severe and Fatigue Dominant long COVID. For example, 25% of SNPs assigned to genes associated with Severe long COVID were included in the top 0.5% of SNPs with the highest feature weights in the ML model for ME. Similarly, 25% of SNPs assigned to genes associated with Fatigue Dominant long COVID were included in the top 2% of SNPs with the highest feature weights in the ML model for ME. These results provide independent validation that a large subset of long COVID genes are also biologically important for ME.

Alternatively, this result could represent an inherent bias in the methodology that would be expected if longer genes mapping to many SNPs are both more likely to be identified by combinatorial analysis and to map to at least one SNP with strong feature importance in the ML model for ME. However, we did not observe the same extreme enrichment of very top ranked model SNPs mapping to endometriosis and coronary artery disease (CAD) disease signatures that we observed for Fatigue Dominant and Severe long COVID. This finding confirms that the very strong enrichment of top ranked ME SNPs that also map to long COVID genes is not an inevitable artifact of any potential gene length biases described above.

Genes mapping to disease signatures for endometriosis and CAD also exhibited significantly higher ranks in the ML model for ME relative to randomly selected genes. This pattern is again apparent when comparing the ML model ranks of the top 25^th^ percentile SNPs for each gene set (Supplementary Figure 7). The degree of overlap is, however, substantially lower than that observed for long COVID, confirming that observation of very strong overlaps between top ranked SNPs in the ML model for ME and Fatigue Dominant and Severe long COVID genes are not an inherent artifact of the analytical approach.

This result implies that some of the mechanisms of action and gene regulatory networks for these two diseases may also overlap with some of those important to ME. Given the broad physiological scope of the mechanisms involved, it is reasonable to expect such overlap, and we have observed evidence for this in combinatorial analyses of these and other diseases (unpublished results).

This gene overlap provides further evidence of the polygenic and heterogeneous nature of ME and long COVID and their partly shared etiology (Taylor *et al*. 2023; Annesley *et al*. 2024). 97 genes (out of 19,146 total annotated autosomal genes) were ranked in the top 20 most-highly ranked genes across at least one of the 10 folds. These 97 genes included 6 genes that were previously associated with long COVID in the Sano Gold cohort (see Supplementary Table 11).

Three overlapping genes (i.e., found in both analyses), *MAPK9*, *NLGN1*, and *PTPRD*, were associated with ‘Fatigue Dominant’ Long COVID, which is significantly more overlap than expected under the null hypothesis (p < 0.001). Each of these genes is associated with a distinct subset of mechanisms previously hypothesized to play an important role in ME and long COVID (Das *et al*. 2022b; Taylor *et al*. 2023; Annesley *et al*. 2024).

*MAPK9*, which encodes mitogen-activated protein kinase 9 (c-Jun N-terminal kinase 2, JNK2), is a stress-responsive kinase that regulates immediate-early gene expression through phosphorylation of key transcription factors (Davis 2000). It integrates inflammatory, oxidative, and metabolic signals—processes relevant to the pathophysiology of both ME and long COVID (Tate *et al*. 2022). Consistent with this, multi-omics analyses in both conditions have identified *MAPK9* as differentially regulated or as a component of disease-associated gene interaction networks (George *et al*. 2022; Lv *et al*. 2022; Peppercorn *et al*. 2023), suggesting a potential role in shared disease biology.

*NLGN1* encodes a postsynaptic adhesion protein critical for glutamatergic synaptic function (Welberg 2012) and is transcriptionally regulated by *CLOCK* (Hannou *et al*. 2018), linking it to circadian rhythm.

*NLGN1* variants and altered expression have been associated with ME (Zhang *et al*. 2025) and with cognitive disorders such as Alzheimer’s disease (Camporesi *et al*. 2021). These findings suggest that *NLGN1* may contribute to shared symptoms of ME and long COVID, including cognitive impairment, sleep disruption, and fatigue, although further studies are needed to confirm this.

*PTPRD*, a receptor-type tyrosine phosphatase, is another neuronal adhesion molecule highly expressed in the brain, where it contributes to synaptic specification and cognitive regulation (Uhl and Martinez 2019). Genetic variants in *PTPRD* have been reported across several human diseases with a neuroimmune component, including Alzheimer’s disease and ME (Uhl and Martinez 2019; Arcos-Burgos *et al*. 2025). Although direct evidence connecting *PTPRD* to long COVID is currently lacking, its neuronal functions and genetic association with ME make it a plausible candidate contributing to the fatigue and cognitive symptoms common across post-viral syndromes.

### Genetic Evidence for Known and Novel Repurposing Opportunities in ME

Our analysis highlighted drug targets with existing therapeutic assets in other indications, enabling systematic identification using a detailed workflow to evaluate potential drug repurposing opportunities in ME based on genetic evidence (Das *et al*. 2022a). For example, we identified novel genetic associations for a target *TLR3* in a patient subgroup that is linked to medications currently under investigation in ME clinical trials (Strayer *et al*. 2012; Mitchell 2016; Strayer, Young and Mitchell 2020).

#### TLR3 and Rintatolimod

Toll-like receptor 3 (*TLR3*) is a key component of the innate immune system that acts as a critical sensor for viral double-stranded RNA in several cell types that are key to host antiviral defense (Vercammen, Staal and Beyaert 2008). Beyond its role in detecting exogenous viral RNA, TLR3 also senses endogenous RNA released by damaged, necrotic, or stressed cells, thereby modulating inflammatory responses (Cavassani *et al*. 2008). Dysregulated TLR3 signaling can lead to chronic inflammation and tissue damage, exacerbating conditions such as autoimmune diseases, chronic viral infections, and cancer (Mohammad Hosseini *et al*. 2015; Hsieh *et al*. 2025)

Rintatolimod is a synthetic double-stranded RNA molecule that acts as a selective agonist of TLR3. On binding to TLR3, rintatolimod activates the MyD88 independent TRIF signaling pathway, leading to the production of interferons and other antiviral proteins without triggering excessive systemic inflammation associated with other dsRNA molecules (Mitchell 2016). It has been investigated in several Phase II/III clinical trials with ME patients, where it has shown statistically significant improvements in primary endpoint using exercise tolerance and some secondary endpoints when compared to placebo (Strayer *et al*. 2012; Mitchell 2016; Strayer, Young and Mitchell 2020).

Despite showing encouraging results and having an acceptable safety profile, rintatolimod still remains an experimental drug in ME in the US, with its use limited to compassionate access programs. The disease signatures associated with *TLR3* identified in a patient subgroup (24.1%) in Analysis 1 could help enhance future trial designs to aid higher efficacy and advance toward full regulatory approval.

In addition, our findings revealed entirely new repurposing opportunities, identifying target–drug pairs involving safe, well-tolerated, generic medications not previously linked to ME. One such example is *PDE4B* and its modulator apremilast, associated with a subgroup of ME patients. Interestingly, our unpublished results suggest that this medication might be also effective in a subgroup of long COVID patients, which reflects shared disease pathology with ME.

#### PDE4B and Apremilast: A Novel Repurposing Opportunity

Our identification of *PDE4B* represents a novel link between this gene and ME. We identified signatures associated with *PDE4B* in around 70% patients in both Analysis 1 and 2. *PDE4B* encodes phosphodiesterase 4B, an enzyme that hydrolyzes cyclic AMP (cAMP), thereby modulating intracellular signaling related to inflammation, immune activation, and cognition (Tibbo and Baillie 2020; Su *et al*. 2022). PDE4B is highly expressed in macrophages, lymphocytes, and CNS glial cells (Jin and Conti 2002; Avila *et al*. 2017), and its dysregulation has been implicated in multiple inflammatory and neurological conditions (Li, Zuo and Tang 2018; Bhat *et al*. 2020).

We hypothesize that overactivation or upregulation of *PDE4B* in ME reduces intracellular cAMP levels, disinhibiting NF-κB and TNF-α signaling pathways (Kida 2025; Ks *et al*. 2025; Zhao *et al*. 2025). This may amplify peripheral and central neuroinflammatory cascades, contributing to core ME symptoms such as fatigue, post-exertional malaise, and cognitive dysfunction. Interestingly, our analysis in long COVID patients identified a modest association with *PDE4D*, another member of the same enzyme family, suggesting a potential shared pathophysiological axis involving dysregulated cAMP signaling (unpublished results).

Among available PDE4B modulators, we prioritized apremilast, an orally available small-molecule PDE4 inhibitor approved for psoriasis and other inflammatory disorders, as the most promising candidate for repurposing. Although non-selective across PDE4 isoforms, apremilast’s inhibition of both PDE4B and PDE4D could confer therapeutic benefits in both ME and long COVID. Its safety profile is favorable, with gastrointestinal side effects such as nausea and diarrhea being the most common adverse events.

Collectively, these findings highlight apremilast as a promising repurposing candidate for ME based on a novel genetic and mechanistic rationale. Future studies should assess its safety and efficacy in mechanistically stratified clinical trials, using biomarkers to identify the subgroup most likely to benefit.

## Discussion

The results of the combinatorial analysis indicate that ME is a highly complex and polygenic disease where genetic factors contribute significantly to increased risk. The findings reinforce and add substantially to the DecodeME GWAS, which identified 8 loci associated with ME in that cohort. We identified 22,411 disease signatures that were significantly associated with increased prevalence of ME in three disjoint patient cohorts. The 7,555 unique SNPs contained in these signatures conservatively mapped to 2,311 genes that are consistently associated with increased ME risk across the three independent cohorts. Choosing a 20% prevalence threshold, the study identified 259 candidate core (i.e., potentially causal) genes, many of which may represent effective drug targets for novel discovery and/or drug repurposing approaches.

These are significant findings with wide reaching implications for study and treatment of the disease.

First and foremost, the results provide further evidence of the clear multisystemic biological basis of ME and suggest a set of candidate core genes and mechanisms that are potentially causal as well as other susceptibility factors. The breadth of genes identified gives multiple new avenues of study, extends the number of drug repurposing candidates that have genetic evidence, and improves the likelihood of finding effective therapies for ME (and long COVID).

### Enabling Targeted Drug Repurposing Trials

Despite ME and long COVID’s massive socioeconomic impact, unmet medical needs, and profound personal and public health consequences (Valdez *et al*. 2019; Al-Aly *et al*. 2024b), there are still no treatments that address the root causes of these diseases.

There are many scientific and technical reasons for this. The biology of the diseases is complex, involving multiple causes and factors, and a wide range of symptoms. They lack a clear, easily measurable biomarker, and there are no reliable animal models that accurately represent the diseases’ pathology. All of these issues add to the uncertainty and risk associated with initiating drug discovery efforts.

The ME community has widely recognized that testing safe and well-tolerated on-market/generic compounds directly in humans through proof-of-concept clinical studies can provide the most rapid and cost-effective route to delivering effective therapies to specific groups of patients. Many such studies are already underway including adaptive trial designs and combination therapy trials (National Library of Medicine 2025; Open Medicine Foundation 2025; Open Medicine Foundation Canada 2025; Ruan et al. 2025). This approach is however constrained by the heterogeneity of the disease, which limits the proportion of responders and the degree of efficacy that can be demonstrated in a non-stratified trial population.

The potential utility of these findings is shown by the association of dozens of genes novel to ME (and long COVID) that include druggable targets. These may be either the basis for new therapeutic programs, or more tractably, using genetically targeted clinical trials of drug repurposing candidates. People with these conditions are often sensitive to medications and adverse effects, and enrolment in trials carries a risk of making their symptoms worse. Better targeted trials might also allow selection only of participants more likely to respond and avoiding those who may be at high risk of worsening of their condition.

This study’s results have the potential to speed up such trials and improve the likelihood of their success. Firstly, the range of candidate core genes identified offers more repurposing targets – a few of these have been highlighted above and there are dozens more potential candidates that could be the subject of future validation studies (Supplementary Materials). Secondly, genetic stratification of the disease offers the potential to identify an individual patient’s likely disease drivers (and resilience factors) and therefore predict which therapies are most likely to be effective for them.

The study used genotype data collected by both UKB and DecodeME in the form of a blood/saliva sample run on a ThermoFisher Axiom UKB genotyping array. This platform could be reused to form the basis of a new non-invasive test that can identify the patients most likely to benefit from a specific intervention during clinical trial recruitment, and also potentially (in the longer-term as part of a new clinical care pathway), the therapies most likely to help an individual patient, once their clinical utility has been established.

Demonstrating effectiveness of a repurposed therapy within a targeted patient cohort would provide essential clinical validation of the disease modifying potential of the novel targets and the ability to select responders. Lack of this evidence is a key risk that prevents biopharma companies from investing further research and development funds in these diseases. Use of genetic/mechanistic biomarkers during drug development to recruit likely responders into clinical trials and build a precision regulatory strategy, could make their clinical development faster and more likely to succeed with fewer adverse events. The same tools may become complementary diagnostics to guide therapy selection in the clinic in due course.

In a similar vein, such a test could be used alongside the lived experience and reported outcomes of patients on studies such as TreatME, which collated thousands of patient’s responses to over 150 treatments (Eckey *et al*. 2025), to further predict a patient’s personal response to a wider range of treatment options.

### Improving Diagnostics and Differential Triage

Outside of the oncology and rare disease space, genetic tests are not usually definitive diagnostics as they describe a person’s lifetime risk of getting a disease rather than their current disease status. In diseases that have environmental or epidemiological components (such as lifestyle or comorbidities) or which are infection mediated, these other factors (e.g. exposure to a triggering infection) play a significant role alongside genetics.

A person’s genetics may however significantly determine their susceptibility and response to infection, their propensity to develop sequelae that persist after the acute phase, which symptoms these may lead to and their severities, and the drugs to which they may respond best (and worst). As predictors of personal risk, and in conjunction with clinical symptoms, genetic tests can therefore be very useful clinical tools, especially in complex diseases, with multiple symptoms that could be mistaken for other conditions or even overlooked.

As shown in this paper, there is a significant overlap of ME disease genes with long COVID, which in some patients may exhibit similar symptoms. Long COVID and ME clearly share multiple genetic commonalities and there are likely to be some drugs that will modulate shared targets, and which may therefore prove to be effective for some patients from both disease groups. Many long COVID genes are, however, not biologically associated with ME and vice versa. These results suggest that they are best considered to be partially overlapping but different diseases.

Similar patterns of significant partial overlap with symptoms and with sets of ME genes have been observed in combinatorial analysis studies of other diseases with a neuroimmune component including fibromyalgia, multiple sclerosis, rheumatoid arthritis, endometriosis, IBD, systemic lupus erythematosus and hypersomnia (unpublished results). These diseases are typically treated by different clinical specialists who may not be able to differentiate the patient’s symptoms accurately.

When fully developed and validated, the insights into genetic similarities and differences between these diseases could be used for rapid non-invasive differential triage of patients presenting with non-specific neuroimmune symptoms. Such tests would facilitate quick and accurate referral of patients to the correct disease specialist to achieve a definitive diagnosis, to evaluate patient prognosis for severity, and to help select the most appropriate therapies for an individual patient.

### Explaining the Genetics of ME

The study’s findings explain significantly more of the disease’s heritable components than have previously been identified, and also why it has historically been so difficult to find or replicate genetic associations for ME using GWAS.

The high observed polygenicity implies that the genetics of ME risk factors are quite variable between individuals, i.e., that every ME patient likely has many genetic variants associated with the disease, but that the sets of genetic variants that are responsible for their personal elevated risk of ME differ between most patients. As discussed, this further implies that understanding the specific sets of variants at play in an individual is going to be crucial for improving diagnostic tests and the success of clinical trials for new or repurposed medicines.

This high polygenic complexity likely also explains why conventional genetic association studies have struggled to identify replicable disease associations. Each additional causal genetic variant effectively acts as a confounding factor that masks the disease association for any single genetic variant, resulting in very small mean effect sizes that are challenging to statistically validate in available datasets.

Nonetheless, the highly complex polygenic basis of ME does not imply that identifying patient subgroups is intractable or that it is not possible to identify effective treatments that apply to large numbers of ME patients. Many of the genes identified in this study show significant enrichment for druggable MoAs such as neurological dysregulation, inflammation, cellular stress responses and calcium signaling (Figure 5) that have been implicated as playing an important role in ME disease biology.

This suggests a commonality in disease biology across patients that is consistent with a quasi-‘omnigenic’ model of disease biology. The omnigenic model of disease hypothesizes that many complex traits such as chronic disease are ultimately directly governed by expression of a relatively small set of ‘core’ genes (Boyle, Li and Pritchard 2017; Ružičková, Hledík and Tkačik 2024; Ratajczak, Heinig and Falter-Braun 2025a). However, identifying those core causal genes via genetic association studies is technically challenging because their expression in the population is directly or indirectly affected by complex gene regulatory networks incorporating a very large number of ‘peripheral’ upstream genes and genetic variants as well as environmental factors (Ratnakumar *et al*. 2020; Ratajczak, Heinig and Falter-Braun 2025b).

A genetic variant mapped to a core gene may increase susceptibility to ME by directly altering expression of that gene, but the effect of that variant in some individuals can be wholly or partially offset by the many other variants throughout the genome that also affect gene expression of the core gene indirectly via altered expression of ‘peripheral’ genes. As a hypothetical, simplified example, assume that a disease state results from overexpression of a single core gene beyond a threshold. In this example, a SNP in a *cis*-regulatory region that directly increases the expression of the core gene will cause some people with the SNP to develop the disease. However, expression of the core gene is also affected by several hormones such as testosterone and estrogen. In some people, levels of circulating hormones decrease the baseline expression of the core gene such that the overall expression remains below the disease threshold despite the increase in expression from the *cis*-regulatory SNP. These people will not develop disease even though they have the ‘causal’ SNP. Likewise, many people who do not possess the *cis-*regulatory SNP could nonetheless develop disease due to hormone-mediated increased expression of the core gene. The genetics of the hypothetical disease would be highly complex as any genetic variant or environmental exposure that directly or indirectly alters levels of circulating hormones could potentially cause expression of the core gene to rise above or drop below the disease threshold. Yet even though it is highly polygenic, it would be simple to effectively treat the disease using therapies that reduce expression of the core gene to levels below the disease threshold.

We hypothesize that ME exhibits similar omnigenic characteristics, and our findings of 259 candidate core genes with an additional set of 2,052 ‘peripheral’ genes that are also reproducible across 3 ME populations is consistent with this view of the disease. This is not an insurmountable complexity problem for study of the disease. It is possible to stratify patients with complex highly polygenic diseases into subtypes characterized by shared disease etiology, e.g., involving misexpression of key metabolic, neurological, or immune response genes, even if the set of peripheral genetic variants driving core causal gene expression differs between patients (Wray *et al*. 2018).

Although the highly polygenic nature of ME suggests that many genes contribute to disease biology, it is possible to gain insights into the core genes and biological processes driving the disease by focusing on the MoAs and cellular pathways associated with the highest prevalence reproducible disease signatures, as well as the signatures and genes with the strongest associations with ME. These can in turn be used to prioritize testing of potential treatments such as drug repurposing candidates aiming to treat ME (and some long COVID) patients.

By representing the subcomponents and non-linear interaction effects of larger gene regulatory networks, combinatorial disease signatures more accurately reflect the omnigenic model of disease biology versus the GWAS and PRS approaches that ignore genetic interactions.

### Moving Towards Improved Prediction of ME Risk

The strong correlation between genetics and ME is notable as genes are likely only one contributing factor towards whether an individual develops ME. Such non-genetic factors are expected to weaken the correlation between genetics and disease status. For example, some ‘high-risk’ controls likely have higher genetic predisposition to developing ME, but may have not been exposed to sufficient triggers, e.g., virus, bacteria, and/or environmental to actually develop the condition. Alternatively, such patients may possess ‘actively protective’ disease signatures which wholly or partially mitigate the effects of their ME disease risk signatures (as described below).

We expect that combinatorial analysis of additional cohorts of ME patients, especially more diverse non-European cohorts, would provide additional insight into the genetics of ME and improve our ability to accurately quantify patient risk. As noted below, actively protective combinatorial analysis is also expected to provide further insights into genetic interactions that govern ME disease biology that are not reflected in the set of double-refined disease risk signatures.

Any future risk score should take into account the hypothesis that ME likely represents several distinct mechanistic subtypes, each with its own disease etiology. For example, a patient might have relatively few disease signatures in total, but a significant excess of signatures associated with a single MoA primarily responsible for their disease subtype. We identified several distinct mechanisms and genetic pathways that were significantly enriched in our disease signatures and were shared across this and all previous combinatorial analyses of ME (and some also with long COVID).

Incorporating mechanistic based stratification into a risk score framework can potentially provide improved insight not only for quantifying an individual’s genetic predisposition towards developing ME but also precision medicine treatments most likely to be effective at treating that patient’s disease.

### Actively Protective Biology and Systematically Identifying Disease Resilience Genes

We can use this deeper understanding of genetic disease risk factors to find people who do not have any form of a disease even though they have a very high level of genetic disease risk factors, have experienced many of the disease triggers, and are as old as possible – so they’ve had every chance to contract the disease and be diagnosed. We can call these people ‘protected’ or ‘resilient’, at least for this specific disease (such protection does not extend to other unrelated diseases).

We have every reason to expect that these protected people should have the disease, so we can speculate that certain biological factors are working behind the scenes to prevent them from showing symptoms. We can find out what makes protected people different by comparing which genes and mechanisms show up significantly more often in them than in diagnosed ME patients. This is an enriched reversal of the usual way we find disease risk genes.

In our UKB ME study, we found many protective signatures, mapping to 9 protein-coding genes that we believe merit further study as they may have a protective effect resisting the onset and progression of ME symptoms (Sardell *et al*. 2025b). Many of these genes’ functions are consistent with alleviating the ME disease risk mechanisms also identified in this study, e.g., in insulin signaling, stress response and autoimmunity. Some of these have also been associated with mechanisms that are subject of approved drugs.

These new ‘actively protective’ or disease resilience genes represent a completely new class of potential drug targets. In the future these may represent opportunities for new or repurposed therapies that could have a prophylactic benefit for many people who have high risk of a specific form of a disease. They could work to reduce the disruption to normal metabolism that causes disease – analogous to statins mitigating cardiovascular risk. Switching on or turning up these beneficial protective mechanisms may well benefit a much wider range of people than just those who have a specific form of the disease.

### Future Studies

The findings presented represent a preliminary combinatorial analysis of the DecodeME dataset. The studies have been performed on just 3 of the 4 batches of DecodeME data and run in just two small case-control cohort studies as opposed to a complete pooled set. There are several other new and confirmatory studies that we would like to perform.

To date neither this study nor the DecodeME study has identified any sex-specific differences between genes associated with ME and we would like to investigate this in more detail.

We have not yet performed actively protective analysis using DecodeME, which we plan to do when the whole dataset has been QCed, and we will also be looking for associations with specific symptoms and/or disease severity.

Finally, formal confirmation that the double-refined signatures are broadly associated with disease (e.g., using the signature count approach) will require an independent cohort of people with ME and controls that are genotyped as part of the same study. This will minimize potential artifacts of batch effects that may produce differences in disease signature frequency between cases and controls that are unrelated to disease biology.

### Limitations of the Analysis

Mis-phenotyping of the cases and controls used due to unreliable or missed diagnosis reduces the number of features detected and the level of reproducibility that can be observed between cohorts (see Supplementary Methods for a fuller explanation). Similarly, we expect genotyping errors, including errors introduced by SNP imputation, would cause reduced reproducibility of signatures, resulting in improper removal or retention of disease signatures and component SNP-genotypes during the Refinement pipeline, as well as reduced predictivity for the set of disease signatures in the Test dataset.

The DecodeME cohort represents the largest and most accurately phenotyped collection of genomic data for ME participants available to date. However, it only contains case samples, and therefore a secondary dataset (UKB) comprised of controls who do not have ME was required to identify disease signatures associated with ME. Although the DecodeME and UKB datasets used for cases and controls used the same Axiom genotyping array (Applied Biosystems 2017), material differences in sample collection and genotyping between the two studies introduced source-specific batch effect artefacts that are inseparable from the case-control phenotype of interest.

With the DecodeME team, we conducted extensive and robust QC procedures to remove all SNPs that showed patterns consistent with batch effects. Only 53-59% of the DecodeME SNPs survived QC and this was further reduced due to lack of overlap between the two sets. However, it is possible that our QC pipeline did not flag all such batch effects, especially those that result in weak disease signal, resulting in potential false positives among the list of identified disease signatures. Conducting secondary screening in UKB mitigates this issue as artefactual signal from lingering batch effects shouldn’t be replicated in the cohort of UKB cases and controls. We believe that any remaining effects are therefore minimal.

However, the highly conservative QC approach used to identify and remove potential batch effects and LD haplotypes may have filtered biologically important SNPs from the dataset as well. We opted to remove any potentially mis-genotyped SNPs as we believed that the potential scientific consequences of Type I error (false positives) in this study are more severe than Type II error (false negatives).

The Refinement pipeline that was used to identify and remove potential false positive SNPs and signatures is also highly conservative and likely prone to Type II error. For example, a SNP-genotype is only retained in a signature if it is associated with increased odds of ME in the full Refinement dataset and all five of the k-fold subsets. However, many signatures are relatively rare and the 95% confidence intervals associated with the observed odds ratios often overlap due to random sampling effects. As such, it is expected that some true biologically important signatures and SNPs will be removed because random sampling variance makes it appear that they are not associated with increased disease risk in one or more of the comparisons. We again elected to accept this high rate of potential Type II error as a trade-off for minimizing potential Type I error and increased confidence in the final set of reproducible signatures and SNPs.

Finally, we applied a conservative gene mapping approach which only assigns SNPs to a gene if they lie within or very close to a gene. This prevents us from identifying any upstream or downstream genetic variants that affect phenotype by altering gene or protein expression. It is likely that these include many of the unannotated SNPs from the double-refined signatures that were not assigned to genes. Alternatively, it is possible that some SNPs located within gene bodies affect phenotype via altered expression of a different gene, or that identified SNPs assigned to one gene are instead in linkage disequilibrium with a true causal SNP mapping to a different gene.

The combinatorial analyses presented in this paper relied exclusively on patients with European ancestry, which despite efforts to the contrary, comprise most (93.1%) of the DecodeME study subjects. This study design was intended to reduce the likelihood of identifying ‘false-positive’ signatures that differ in frequency between cases and controls not due to disease biology but because they occur at different background frequencies in populations with differing rates of ME diagnosis. Populations with other ancestries were not sufficiently well-represented to enable a statistically robust analysis.

In the omnigenic disease model, effect sizes of individual SNPs are expected to vary between groups with different ancestries due to population-level differences in the frequency of confounding peripheral genetic variants, which has been proposed to explain the poor predictive performance of polygenic risk scores across ancestries (Mathieson 2021). Combinatorial analyses of non-European ancestry populations may therefore provide insight into novel genes and disease signatures that are associated with ME in all populations but that have relatively greater importance in some patient subgroups due to the complex combinatorial nature of ME.

We note, however, that previous combinatorial analyses of long COVID and endometriosis disease signatures have demonstrated broad reproducibility (51-94%) of combinatorial signatures and genes derived from European populations across patient cohorts with diverse ancestries (Sardell, Das, Møller, et al., 2025; Sardell, Pearson, et al., 2025), suggesting that many of the insights of this study are likely to be applicable to non-European patients as well. In part this high reproducibility results from the disease signatures being comprised of combinations of relatively common variants that are represented more consistently across multiple ancestries than rarer variants.

## Conclusions

The findings of this study substantially increase the number of genetic associations with ME, providing further evidence that ME is a complex multisystemic disease. This is an important step forward in scientific and clinical understanding of the disease and for the patient community who have been overlooked for so long.

ME has been shown to be a highly polygenic and heterogenous disease, with a set of over 250 candidate core disease risk genes across a variety of mechanisms. This has significant implications for research strategy and the opportunity to develop more personalized clinical pathways. Stratification of patients by the mechanisms driving their form of the disease will be critical to predicting which patients will benefit from which therapy. It will also provide a crucial tool for developing more accurate diagnostic tests, creating patient selection tools that can make clinical trials more successful, and identifying novel and repurposed drugs.

These results confirm the findings from other DecodeME based studies and its role as a key enabler of research into ME. While not without challenges as a dataset, the utility of DecodeME and combinatorial analytics as a research tool for ME and long COVID has been clearly demonstrated, and we hope this will encourage the development of new data collections and studies with more participants, including from diverse ancestries, with more detailed longitudinal clinical histories and deeper multiomic characterization of participants across multiple time points.

These are, however, only preliminary findings and there are several on-going analyses to answer more specific questions. We are now beginning to uncover ME targets and tools with the precision required for more coordinated drug research. We hope that this will stimulate research across the community and encourage people with ME and long COVID to continue to contribute their samples, data and time to future studies.

## Data Availability

Only data from existing DecodeME and UK Biobank study cohorts were analyzed and no new source data were collected for this study. DecodeME data can be accessed by approved studies (https://institute-genetics-cancer.ed.ac.uk/decodeme-the-worlds-largest-mecfs-study/researcher-access) and UK Biobank data can be accessed by approved registered users (https://www.ukbiobank.ac.uk/about-us/how-we-work/access-to-uk-biobank-data/).
PrecisionLife will make the results of its analysis available and will support bona fide researchers in testing the drug repurposing candidates identified.

## Availability of Data and Materials

Only data from existing DecodeME and UK Biobank study cohorts were analyzed and no new source data were collected for this study. DecodeME data can be accessed by approved studies (https://institute-genetics-cancer.ed.ac.uk/decodeme-the-worlds-largest-mecfs-study/researcher-access) and UK Biobank data can be accessed by approved registered users (https://www.ukbiobank.ac.uk/about-us/how-we-work/access-to-uk-biobank-data/).

PrecisionLife will make the results of its analysis available and will support *bona fide* researchers in testing the drug repurposing candidates identified.

## Author Contributions

JMS, SG, MAS and SD contributed to the design of the study. JMS designed the analytic pipelines including signature Discovery, Refinement and signature count score testing. JMS, SG, SD, ARM and MAS wrote the manuscript. MP and MSperformed dataset generation and QC. MP performed batch effect analysis and combinatorial analysis of datasets. SD performed analysis of genes and overlap with other studies. ARM performed analysis of genes and potential repurposing candidates. DK performed imputation and machine learning analysis on overlap of ME and long COVID. LMF performed signature Refinement. AR supported related clinical studies and data access. HB, KM, MN, DL, SC, and AR provided input from people with lived experience input and feedback throughout the project. All authors provided input and approved the final version of the manuscript.

## Acknowledgements

We thank Chris Ponting, Sjoerd Beentjes, Esther Ewaoluwagbemiga and the DecodeME team, the Action for ME team, Amy Rochlin and the CODA team, Oved Amitay, María Paz Muñoz, Rowan Gardner, Rohit Gupta, Mark Bouzyk, and Lynn Viatge for their active participation, support and invaluable feedback through the research process. Thanks to James Kozubek and Julie Borgel for preliminary analysis, along with Sonia Luczynska and Alexis Elves for coordination of LOCOME project meetings and other administrative tasks. Special thanks to Gert Møller and Claus Erik Jensen, who initially developed the combinatorial analytics methodology, and the rest of the PrecisionLife team who provided support for the project.

We gratefully acknowledge and thank the DecodeME, Sano GOLD, and UK Biobank participants, and the wider network of patients with lived experience of the disease for their contributions without whom this research would not have been possible. This work uses data provided by patients and collected by the NHS as part of their care and support, for which we are also grateful.

Research described in this article has been conducted using data from the DecodeME and UK Biobank (application number 44288) studies. We also thank DecodeME team, which includes Action for M.E. and the MRC Human Genetics Unit, University of Edinburgh and UK Biobank, for making available the participant data examined in this study.

## Ethics Approval and Consent to Participate

Research described in this article has been conducted using data from DecodeME study and UK Biobank (application number 44288).

The DecodeME study has approval from North West - Liverpool Central Research Ethics Committee (21/NW/0169). DecodeME is registered in the Research Registry (identifying number 6395). More information on the study is available online (www.decodeme.org).

UK Biobank has approval from the North West Multi-centre Research Ethics Committee (MREC) as a Research Tissue Bank (RTB) approval, and researchers do not require separate ethical clearance and can operate under this RTB approval.

