## Supplementary Methods & Data for "Identification of Novel Reproducible Combinatorial Genetic Risk Factors for Myalgic Encephalomyelitis in the DecodeME Patient Cohort and Commonalities with Long COVID"

#### Basic DecodeME Dataset QC

We first filtered the variants to include only bi-allelic SNPs that overlapped with our dataset of potential controls from UKB. We then filtered by missingness (removing all individuals with >2% of SNPs missing and all SNPs that are missing from >2% of samples). Additional QC included filtering SNPs that significantly deviate from Hardy-Weinberg Equilibrium ($p < 10^{-12}$), removing SNPs with minor allele frequency < 0.01, and filtering participants with close relatedness. Finally, we removed all SNPs independently flagged by the DecodeME project team as having low quality genotype calls during QC (DecodeME, 2025). Participants were assigned to ancestral populations using NCBI's GRAF-pop tool (Jin *et al.* 2017), and any participants not assigned to GRAF-pop's "white" subgroup were removed.

After QC, we retained 12,753 cases across the four DecodeME cohorts (Table 1). The fraction of participants flagged as non-European and removed by GRAF-pop (6.9%) is significantly larger than the percentage of participants with self-reported European ancestry in the GWAS study design (3.5%) (DecodeME 2025). This is likely in part due to the relatively low number of ancestry-specific genetic markers employed by GRAF-pop that are included on the UKB Axiom array and survived QC in our datasets. Reduced sampling of ancestry specific markers can cause GRAF-pop to incorrectly assign some participants with European ancestry to 'Hispanic' or 'other' populations, while maintaining high population assignment accuracy among patients flagged as 'European' (Jin *et al.* 2019). Use of GRAF-pop may have led to removal of some case samples that could have been used, but the loss of statistical power associated with fewer samples is preferable to less conservative options which can potentially lead to identification of signatures not directly linked to disease.

#### Identification and Removal of Batch Effects

Initial QC removal of batch effects focused on DecodeME Batch 1 (Cohort F), which was the first set of DecodeME patient data available for analysis, merged with UKB Discovery controls (Cohort B) as described in Materials and Methods. We first ran GWAS on the merged dataset using PLINK 1.9 (Purcell *et al.* 2007)and removed any SNPs that achieved statistical significance at a stringent threshold of $p < 1x10^{-8}$.

The logic behind this decision was that previous GWAS studies in similar sized datasets found no replicable significant single-SNP genetic associations with ME (Dibble, McGrath and Ponting 2020), and therefore any strong associations were more likely to represent batch effects from merging. Nearby SNPs that should be in linkage disequilibrium (LD) to the significant SNPs did not show strong associations with disease, resulting in absence of the multi-SNP peaks in the GWAS Manhattan plot that would be expected for true SNP-diseases associations. Such patterns strongly imply batch-specific genotyping errors affecting a single SNP. We further confirmed that none of the identified outlier SNPs were associated with strong effect sizes in a GWAS for the UKB cohort described below (Cohort C+E), consistent with them being likely batch effects.

Combinatorial analysis is an iterative process and can therefore be more sensitive than GWAS to relatively rare genotyping errors that are restricted to either cases or controls. Such rare errors often have relatively small impacts on GWAS summary statistics and do not result in significant deviation from Hardy-Weinberg equilibrium, so they cannot be detected using conventional methods for filtering batch effects. However, in a combinatorial analysis, they manifest as rare multi-SNP haplotypes that have very strong odds ratios and highly significant *p*-values of association (Lee *et al.* 2010).

For example, consider a scenario in which 5% of cases and 0% of controls are improperly assigned a heterozygous ('1') genotype due to incorrect genotype calling of the SNP in a sample of cases. This SNP is in perfect linkage disequilibrium (LD) with a nearby SNP B in the genome, such that the minor allele of the former is always co-associated with the minor allele of the latter in the broad population. However, the batch-specific genotyping error results in 5% of cases who are improperly assigned the non-existing haplotype A1-B0, which in turn is strongly and significantly associated with the disease phenotype. Combinatorial analysis is intended to identify such relatively rare combinations of SNP-genotypes with strong effect sizes, so it is important to ensure that these are associated with true biological signal.

After applying the PrecisionLife combinatorial analytics platform to the initial Discovery dataset comprised of DecodeME Batch 1 cases (Cohort F) merged with UKB Discovery controls (Cohort B), we identified over 60,000 signatures that occurred at extremely elevated frequency in cases relative to controls. All signatures contained at least two SNPs that were co-located in the same genetic region ('LD haplotypes') consistent with the abnormal LD patterns that are expected to arise from batch-specific misgenotyping (Lee *et al.* 2010). We also observed a small set of SNPs that occurred at high frequency in the signatures, often found in combination with different co-located SNPs. Again, none of these extreme disease associations were present in the UKB cohort.

For each LD haplotype, we filtered the SNP with the strongest odds ratio in the PLINK GWAS described above, under the assumption that it is more likely to represent the misgenotyped variant in a pair. In this manner, we identified and removed 161 suspected 'batch-effect' SNPs. Although we did not have access to genotyping clustering data, the DecodeME team confirmed via manual inspection of genotyping plots that a sample of SNPs selected in this manner all exhibited problematic patterns of genotype clustering assignments, suggesting that the approach is effective at identifying and removing genotyping error batch effects.

Next, we ran a combinatorial analysis on the filtered dataset and again detected disease signatures containing novel LD haplotypes that had disease associations that were lower than those observed in the initial analysis but still highly elevated.

Because all possible combinations of SNPs cannot be evaluated, the PrecisionLife combinatorial analytics platform identifies the optimal search space with the strongest likelihood of identifying significant disease associations. Removing the initial set of mis-genotyped SNPs allowed the platform's search algorithm to test novel combinatorial signatures featuring these weaker batch effects. We identified and removed 56 additional likely mis-genotyped SNPs from LD haplotypes as described above. This process was repeated until the combinatorial analysis returned zero signatures that contained a pair of SNPs from the same genomic locus.

We applied the same QC pipeline to DecodeME Batches 2+3 (Cohort G+H), identifying and removing an additional 16 potentially mis-genotyped SNPs based on the presence of haplotype signatures in the output of the combinatorial analysis. For these batches, we also removed any SNPs not included in the QCed merged Batch 1 dataset (Cohort B+F).

To identify novel batch effects, we also conducted pairwise batch vs. batch analyses using both GWAS and combinatorial analytics. First, we conducted a PLINK GWAS with batch number as the 'case-control' phenotype of interest and removed any GWAS-significant SNPs ($p < 5x10^{-8}$). We then applied the same iterative approach described above to flag and remove any SNPs that resulted in identification of disease signatures in a batch vs. batch combinatorial analysis. In this manner, we identified and removed a further 739 SNPs from all DecodeME cohorts that were likely associated with DecodeME batch-specific artefacts.

After pooling and randomly re-splitting cases from DecodeME Batches 2 & 3 (Cohort G) and merging with UKB Discovery controls (Cohort B) as described above, we again conducted combinatorial analysis on each split to confirm that we had successfully removed potential batch effects from the data.

Manhattan plots of the Analysis 1 Discovery, Refinement, and Test DecodeME datasets are shown in Supplementary Figures 1-3.

#### Dataset Imputation

Over 14% of the SNPs present in the Analysis 1 Discovery dataset (Cohort B+F) were not present in either the Analysis 1 Refinement (Cohort C+G) or Test (Cohort D+H) datasets due to filtering of poor-quality SNP calls either during the original genotyping or removal of batch effects during this study. As a result, 46% of the identified disease signatures could not be refined or validated using available genotype data because they contained at least one of these missing SNPs.

Imputation of the missing SNP genotypes was performed using Beagle v5.4 (27Feb25.75f) (Browning, Zhou and Browning 2018) on a merged DecodeME dataset comprised of Batches 1 through 3 (Case Cohorts F+G+H and Control Cohorts B+C+D). To manage computational constraints, the dataset was split into three random subsets containing participants from each batch and further divided into per-chromosome VCF files for chromosomes 1-22. Post-imputation processing included conversion to PLINK2 binary format, filtering variants based on their presence in DecodeME, removing duplicates, and merging subsets. The final imputed dataset was resplit into original batch structures while preserving sample order and phenotype information for downstream genetic association analyses.

We did not perform imputation on the Discovery datasets due to the compounding nature of genotyping errors in a combinatorial context. For example, a mean SNP genotyping error of 2% implies that nearly 10% of participants will be improperly assigned with respect to a five-SNP disease signature. Such errors can result in reduced efficiency of the platform's heuristic search algorithm's ability to identify disease signatures.

#### Calculation of Genetic PCs

Genetic PCs were calculated for the validation dataset using principal component analysis (PCA) implemented in PLINK 1.9. Before calculating PCs, we first performed LD-pruning using the command `--indep-pairwise 50 5 0.2`. We then removed sex-linked SNPs, SNPs with minor allele frequency less than 0.05, and SNPs located within the low recombination MHC region (chr6:28,477,797-33,448,354). We opted not to include more PCs to avoid spurious results that can result from including many PCs in the analysis (Grinde *et al.* 2024).

#### Analytical Pipeline – Signature Refinement

We first refined the output of the combinatorial analyses using a dataset comprised of 1,985 self-reported ME cases from UKB's Pain Questionnaire (Cohort E) and 112,824 UKB controls (Cohort C). This case cohort, which was previously used to identify disease signatures, is described in more detail in Das et al. (2022). Although phenotyping in UKB is not as robust as DecodeME, this step allows us to remove potential batch effects from merging cases and controls from different datasets as they will not be reflected in disease associations in the unmerged UKB cohort.

The Refinement process first entailed running a 'strong components' test that identifies and removes SNP-genotypes within a disease signature if they do not improve the signature's association with risk. That is, for each signature, we only retained a component SNP-genotype if removing it resulted in a signature with a weaker odds ratio in the Refinement dataset. We conducted this analysis in the full UKB Refinement cohort and using a 5-fold subsampling approach where we removed 20% of the samples to ensure repeated robustness of the strong components results. Only SNP-genotypes that were repeatedly associated with increased risk across all 5 folds of the test were retained, outputting a list of 'strong component' disease signatures.

Next, we removed all strong component disease signatures that had an odds ratio less than 1 in the UKB cohort (Cohort C+E). Some strong component signatures were comprised of a set of SNP-genotypes that were also incorporated within at least one higher-layer signature (e.g., the signature *A-B-C* is contained within the signature *A-B-C-D*). We identified all such pairs and tested whether participants with the lower-layer signature but not higher-layer signatures (e.g., the signature *A-B-C* but not *A-B-C-D*) have odds ratios less than or equal to 1. If so, then we considered the lower-layer signature to be 'redundant' to the higher-layer signature and removed the former from the list of refined signatures.

Finally, we 'expanded' the set of refined signatures to capture alternative genotypes that are also associated with disease. For example, the combinatorial analytics platform may only report signatures that have a heterozygous ('1') genotype at a causal SNP even though the homozygous minor allele ('2') genotype has a stronger effect size (i.e., odds ratio). This reflects the greater statistical power for validating disease associations for the more common 1 genotype signatures relative to the rare 2 genotype signatures. Alternatively, the platform may only report signatures with a 2 genotype but not signatures with a 1 genotype if the latter have smaller but still biologically relevant effect sizes relative to the 0 genotype. Incorporating these biologically important alternative genotypes into the set of signatures allows us to capture a broader spectrum of disease biology.

To expand the set of signatures we first evaluated a 'dominant' genetic model which substituted 1 genotypes for 2 genotypes in the refined signatures and vice versa and then evaluated the odds ratios of the expanded signatures in both the original Discovery dataset (Cohort B+F or B+G) and the UKB Refinement dataset (Cohort C+E). Expanded disease signatures with odds ratios greater than 1 in both datasets were added to the set of refined disease signatures. We then did a similar analysis for a 'recessive' genetic model where 0 genotypes were substituted for 1 genotypes and vice versa.

Next, we took the set of signatures that passed the Refinement pipeline in UKB and applied an identical Refinement approach using the appropriate DecodeME Refinement dataset (Cohort G+C or F+C). Finally, we checked that the set of signatures resulting from Refinement in UKB and DecodeME were still associated with ME (odds ratio > 1) in the corresponding Discovery dataset (Cohort B+F or B+G) and removed any that were not.

#### Machine Learning Model for ME

We first trained a heavily-regularized XGBoost model using 10-fold cross validation to predict ME case/control status in in a merged dataset of 10,569 cases from DecodeME Batches 1-3 (Case Cohorts F+G+H) and 21,138 UKB controls (for a 1:2 case:control ratio), including 587,174 SNPs (after SNP imputation). The model AUROC was 0.96 on the Train set and 0.57 on the Test set (mean over 10 folds).

Within each of the 10 folds we obtained the feature importances of every SNP for each sample in the test set and calculated the mean feature importance for each SNP across samples. We then ranked SNPs in each fold by mean feature importance and calculated its median ranking across the 10 folds. SNPs were mapped to a list of 19,146 autosomal protein-coding genes using a conservative approach: a variant was assigned to a gene if it is located within its cis-region (gene borders with 1kbp padding on each side). All variants that are not located within a cis-region of any protein-coding gene were assigned a 'dummy gene' annotation.

We conducted two analyses to test whether the genes associated with long COVID are also assigned high feature importances in the ML model for ME.

First, we tested whether genes associated with each long COVID cohort are broadly enriched in the set of top ranked SNPs in the ML model for ME. For each gene set, we identified the median and top 25$^{th}$ percentile ranks of all SNPs in the ML model for ME mapping to those genes (where top 25$^{th}$ percentile rank corresponds to the SNPs with the top 25% feature weights). We randomly selected the same number of genes from the full set of protein-coding genes and calculated the median and top 25$^{th}$ percentile rank for SNPs mapping to the random set of genes. We repeated that process 10,000 times to identify a distribution of median and top 25$^{th}$ percentile ranks under the null hypothesis. We used this to assign *p*-values of randomly observing median and top 25$^{th}$ percentile ranks comparable to the observed values for each gene set.

For comparison, we conducted similar enrichment analyses using SNPs mapping to 1097 genes found in a UKB combinatorial analyses of endometriosis patients and 133 genes found in a UKB combinatorial analysis of coronary artery disease. The dataset and output of the combinatorial analysis for endometriosis are described in Sardell et al. (2025). For coronary artery disease, we conducted a combinatorial analysis of 499,639 SNPs for 13,435 UKB cases and 26,865 UKB controls who satisfy the criteria listed in Supplementary Table 10.

We also identified key long COVID genes that also were very strongly linked to ME in the ML model. For each fold, we identified the set of 'high-feature-importance' ME genes corresponding to the top 20 ranked SNPs, after excluding SNPs assigned to the 'dummy gene' category. We then pooled the sets of high-feature importance genes across all folds and checked for overlap with the genes associated with each long COVID cohort.

For each gene overlap result, we again used a permutation-based approach to assign *p*-values. First, we randomly selected *k* genes from the full set of protein-coding genes, where *k* is the number of genes associated with the long COVID cohort. We then identified how many of those random genes overlapped with the set of high-feature importance genes from the ML model for ME.

This approach was again repeated 10,000 times to generate a distribution of gene overlap under the null hypothesis and the *p*-value is equal to the percent of permutations in which *k* or more overlapping genes were identified. We opted to apply a permutation-based approach to assign *p*-values rather than a binomial likelihood calculation because genes have unequal probabilities of being assigned a top rank in the ML model due to different numbers of SNPs per gene represented in the QCed genotype data.

#### Length-matched Permutation Test for Long COVID Gene Overlap

We performed an enrichment analysis to assess the overlap between double validated signatures and long COVID genes using a length-matched permutation test to account for potential gene length bias. Approximately 20,000 protein coding genes were used as background gene set which was partitioned into three bins of based on gene lengths defined by length cutoffs at 50 and 250 kb: [0,50), [50, 250), and ≥ 250 kb.

Random gene sets of exactly the same size as the set of ME genes mapped to double-validated signatures were drawn from the background gene set in 10,000 iterations to match the gene length distribution of each bin, thereby generating an empirical, length-adjusted null distribution. *p*-values were calculated for each gene length bin to assess enrichment of long COVID gene overlap with ME genes in comparison to the length-matched random gene permutations.

### Supplementary Results

#### Signature Discovery and Refinement

As explained in the main text, we conducted two complementary analyses of the DecodeME dataset, swapping the cohorts of participants used for Discovery and Refinement.

##### *Analysis 1*

We initially identified 150,683 disease signatures for ME comprised of 4,209 unique autosomal SNPs in our initial combinatorial analysis of the Analysis 1 Discovery dataset (Cohort B+F) (Supplementary Table 3).

Applying the signature Refinement pipeline to the set of initial signatures returned 43,816 signatures comprised of 3,679 SNPs that were consistently associated with increased odds of ME in the UKB Refinement dataset (Cohort C+E) ('UKB-Refined signatures') (Supplementary Table 3). The substantial reduction in the number of signatures after applying the Refinement pipeline does not imply that the disease signatures output by the combinatorial analysis fail to replicate in bulk. A key aim of this Refinement step is to identify the core set of most reproducible SNPs and multi-SNP combinations that are shared across disease signatures. As a result, each Refined signature typically corresponds to multiple signatures from the original output.

Further refinement in the Analysis 1 DecodeME Refinement dataset (Cohort C+G) resulted in a set of 12,463 signatures, comprised of 2,748 SNPs mapped to 1,001 protein-coding genes, that were consistently associated with increased odds of ME in multiple DecodeME cohorts and UKB (see Supplementary Table 4 for the number of SNPs in the combinations). We refer to these as 'double-refined signatures'.

Ignoring genotypes, these signatures represent 11,463 combinations of SNPs. 1,576 signatures were removed from the analysis during this Refinement step because they were missing at least one SNP that could not be reliably imputed in the Analysis 1 DecodeME Refinement dataset (Cohort C+G). 31% of the double-refined Analysis 1 signatures have odds ratios greater than 1.1 in the Analysis 1 Refinement dataset, with a maximum odds ratio = 2.22 and mean and median odds ratios = 1.09 (Supplementary Figure 4).

##### *Analysis 2*

Initial combinatorial analysis of the Analysis 2 Discovery dataset (Cohort B+G) identified 95,916 signatures comprised of 8,101 unique SNPs (Supplementary Table 3). This represents fewer signatures but more SNPs than the combinatorial analysis for Analysis 1 even though the Analysis 2 Discovery dataset included nearly 15% fewer SNPs than the Analysis 1 Discovery dataset.

Refining the Analysis 2 signatures in the UKB Refinement (Cohort C+E) and the DecodeME Analysis 2 Refinement (Cohort C+F) datasets output 9,963 double-refined signatures (Supplementary Table 3), comprised of 5,008 unique SNPs, mapped to 1,647 protein-coding genes that are consistently associated with increased odds of ME in multiple DecodeME cohorts and UKB.

Ignoring genotypes, these signatures represent 9,048 combinations of SNPs. 39% of the double-refined Analysis 2 signatures have odds ratios greater than 1.1 in the DecodeME Analysis 2 Refinement dataset (Cohort C+F), with a maximum odds ratio = 2.55, mean odds ratio = 1.10, and median odds ratio = 1.11 (Supplementary Figure 4).

#### *Overlap between Analysis 1 and Analysis 2*

The double-refined signatures from Analysis 1 and Analysis 2 shared 201 SNPs, including 15 identical signatures common to both. In total, 337 genes map to at least one double-refined signature from both analyses, 76 of which mapped to shared SNPs. We did not observe any significant differences when comparing the odds ratios of the shared signatures or SNPs to signatures and SNPs that were identified in one analysis.

We hypothesize that the relative lack of overlap between the results of Analysis 1 and Analysis 2 primarily reflects a combination of the relatively limited sizes of the Discovery datasets and the highly complex disease biology of ME/CFS. Under the omnigenic disease model the subcomponents of the larger gene regulatory network that are most important for disease vary by patient and by cohort based on the presence/absence of genetic and environmental confounders.

This has been hypothesized to explain the well characterized pattern in which the effect sizes (i.e., odds ratios) of SNPs for complex chronic diseases in a GWAS context vary considerably between different cohorts (Mathieson 2021). That is, SNPs are generally consistently positively associated with disease, but the relative ranks of SNPs often vary widely across cohorts except for SNPs with the strongest effect sizes. This inconsistency is magnified when datasets are small and the observed effect sizes for a sample are more likely to randomly deviate from the mean effect sizes in the larger population. Indeed, the GWAS Manhattan plots for the Analysis 1 Discovery, Refinement, and Test datasets exhibit little overlap in terms of SNP effect sizes (Supplementary Figures 1-3).

The variance in effect sizes for SNPs across cohorts has important consequences for overlap between the output of two combinatorial analyses. When identifying disease signatures, it is only computationally feasible to test a tiny fraction of the total possible combinations of SNP genotypes. The PrecisionLife platform overcomes this challenge by employing a deterministic heuristic algorithm to identify the ‘combinatorial search space’ which offers the greatest potential for identifying significant disease associations during each layer. If the relative ranks of effect sizes for SNPs vary substantially between cohorts, then the optimal combinatorial search space identified by the disease signature mining algorithm during each run can be quite different, resulting in different sets of SNPs and genes.

Lack of overlap between combinatorial analysis can also reflect the common pattern from GWAS in which effect sizes of significant SNPs are inflated in the original cohort relative to replication cohorts, a phenomenon called “winner’s curse” (Forde, Hemani and Ferguson 2023).

Because genetic associations rely on a small sample of the larger population, the observed effect size of a genetic feature will almost never be exactly equal to the true biological effect size – sometimes the observed feature will be randomly over-represented in cases (resulting in an observed effect size larger than the true effect size) and sometimes it will be randomly over-represented in controls (resulting in an observed effect size smaller than the true effect size). Applying a minimum *p*-value threshold favors features with effect sizes that have been the most strongly inflated by random sampling effects. When the effect sizes of these SNPs are later measured in a second dataset, the observed effect size is expected to continue to reflect the biological association of the SNP with disease but without the same inflation due to random sampling.

This same dynamic also affects the disease signature mining algorithm in a combinatorial analysis – the mining algorithm favors features that are simultaneously biologically associated with disease and randomly over-represented in cases in the sample relative to the larger population. The former is expected to replicate across cohorts but the subset of biologically important features that are also subject to random effect size inflation will not, resulting in different 'optimal' search spaces between runs.

If the number of genes biologically related to disease is small (or if a small subset of genes have strong effect sizes), then the combinatorial search spaces explored by the mining algorithm are expected to be similar across cohorts resulting in high overlap in output. But when the number of genes related to disease is very large and the effect sizes of features are similar, as these results suggest for ME, the relative importance of genes is expected to vary randomly between cohorts increasing the likelihood that combinatorial analyses will identify different sets of features. This is especially true in relatively small datasets, as the statistical likelihood of encountering relatively large differences between the observed and true effect sizes is inversely correlated with dataset size.

Importantly, lack of overlap between the output of two combinatorial analyses does not imply that the disease signatures identified in one cohort are not associated with disease in the second cohort - merely that they are expected to have lower effect sizes in the second cohort relative to the disease signatures that were identified in that cohort. The refinement process ensures that signatures and SNPs are correlated with disease in both cohorts even if they were only identified in one combinatorial analysis.

The predictive performance of the pooled set of double-refined disease signatures derived from Analysis 1 and Analysis 2 is significantly better than the predictive performance using either set of signatures individually. This demonstrates that both analyses provide important and novel insight into the biology of ME, and that a planned analysis of the full pooled DecodeME population (when available) may generate further associations.

#### Limitations of the Analysis

Genetic association studies entail comparing DecodeME participants (cases) to a secondary dataset (in this case UKB) comprised of healthy controls who do not have ME. However, the often infection mediated nature of ME poses challenges for all genetic association studies as some samples included in the healthy controls likely have high genetic susceptibility to developing ME, but have not been exposed to a triggering infection or other causal factor that might have led to ME, or they may have substantial protective genes despite facing these triggers. ME's highly polygenic and heterogenous nature also poses challenges both due to missed and misdiagnosis. ME cases in UKB were identified based on self-reported non-disease specific survey response data, which can improperly conflate patients with ME with patients affected by chronic fatigue unrelated to ME.

Such mis-phenotyping in both cases and controls works to reduce the difference between cases and controls and may potentially result in improper removal of biologically relevant signatures and SNPs during the Refinement process. Mis-phenotyping therefore reduces the number of features detected and the level of reproducibility that can be observed between cohorts.

The highly conservative QC approach used to identify and remove potential batch effects may have filtered biologically important SNPs from the dataset as well. Notably, we flagged any linkage disequilibrium (LD) haplotype that occurred in an identified disease signature as a potential artefact of batch effects, which removed 900 SNPs from the analysis. However, we have previously identified multiple disease signatures containing LD haplotypes when conducting combinatorial analyses of other diseases where cases and controls were drawn from the same dataset and therefore shouldn't be subject to strong batch effects (Taylor *et al.* 2023; e.g., Sardell *et al.* 2025).

LD haplotypes can be correlated with disease when they represent pairs of SNPs subject to intra-locus epistasis, e.g., between a SNP that alters a protein's structure and one that modifies expression of the protein-coding gene. Alternatively, LD haplotypes can be correlated with disease when they act as reliable proxies for nearby causal variants that are absent from the dataset.

Unfortunately, the perfect correlation between case-control status and dataset source in the source dataset prevents us from distinguishing LD haplotype signatures that are artefacts of batch effects from LD haplotype signatures that reflect disease biology. We therefore opted to remove any potentially mis-genotyped SNPs as we believed that the potential scientific consequences of Type I error (false positives) in this study are more severe than Type II error (false negatives).

SNP imputation introduces another potential source of error in the Refinement analysis. Unfortunately, this was necessary to evaluate the full set of disease signatures identified in Analysis 1 as many Batch 1 SNPs (Case Cohort F) were missing from the DecodeME Batch 2+3 datasets (Case Cohorts G and H). Some of these missing SNPs were filtered during batch-specific QC conducted as part of this study. However, many were absent from the original DecodeME genotyping data presumably due to removal of poorly genotyped SNPs in the DecodeME SNP calling pipeline. In general, we expect that genotyping errors should cause reduced reproducibility of signatures, resulting in improper removal or retention of disease signatures and component SNP-genotypes during the Refinement pipeline, as well as reduced predictivity for the set of disease signatures in the Test dataset.

### Supplementary Figures

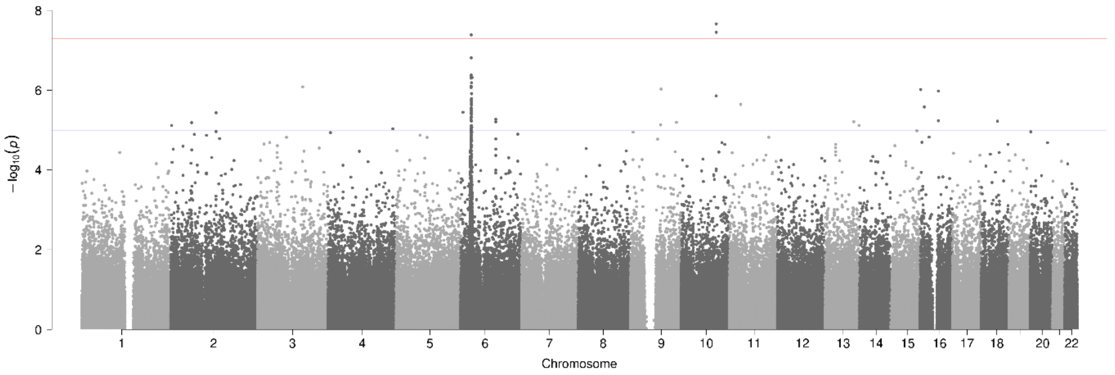


**Supplementary Figure 1.** Manhattan plot generated using PLINK of genome-wide *p*-values of association for the Analysis 1 Discovery dataset comprised of 3,405 cases from DecodeME Batch 1 (Cohort F), 38,903 controls for UKB (Cohort B) and 408,873 SNPs. The horizontal blue and red lines represent the genome-wide significance values of $p < 1x10^{-5}$ and $p < 5x10^{-8}$ respectively.

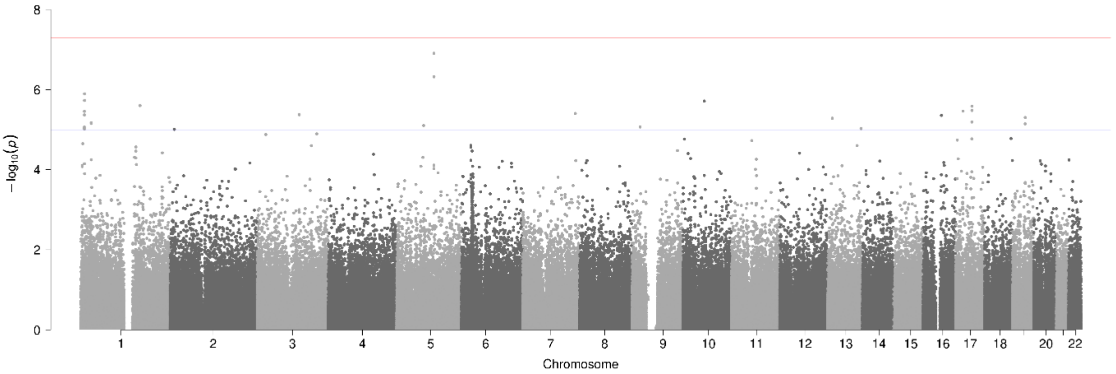


**Supplementary Figure 2.** Manhattan plot generated using PLINK of genome-wide *p*-values of association for the Analysis 1 Refinement dataset comprised of 3,585 cases from DecodeME Batches 2+3 (Cohort G), 107,409 UKB controls (Cohort C) and 349,734 SNPs. The horizontal blue and red lines represent the genome-wide significance values of $p < 1x10^{-5}$ and $p < 5x10^{-8}$ respectively.

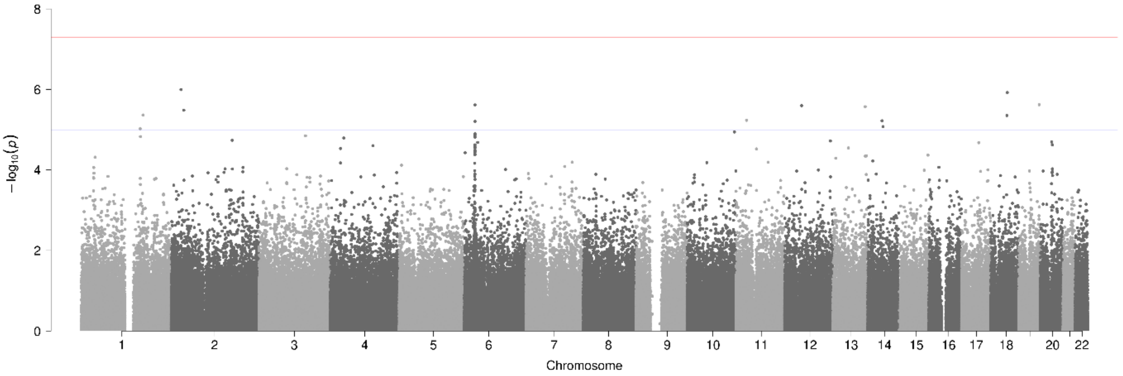


**Supplementary Figure 3.** Manhattan plot generated using PLINK of genome-wide *p*-values of association for the Test dataset comprised of 3,579 cases from DecodeME Batches 2+3 (Cohort H), 107,619 UKB controls (Cohort D) and 350,014 SNPs. The horizontal blue and red lines represent the genome-wide significance values of $p < 1x10^{-5}$ and $p < 5x10^{-8}$ respectively.

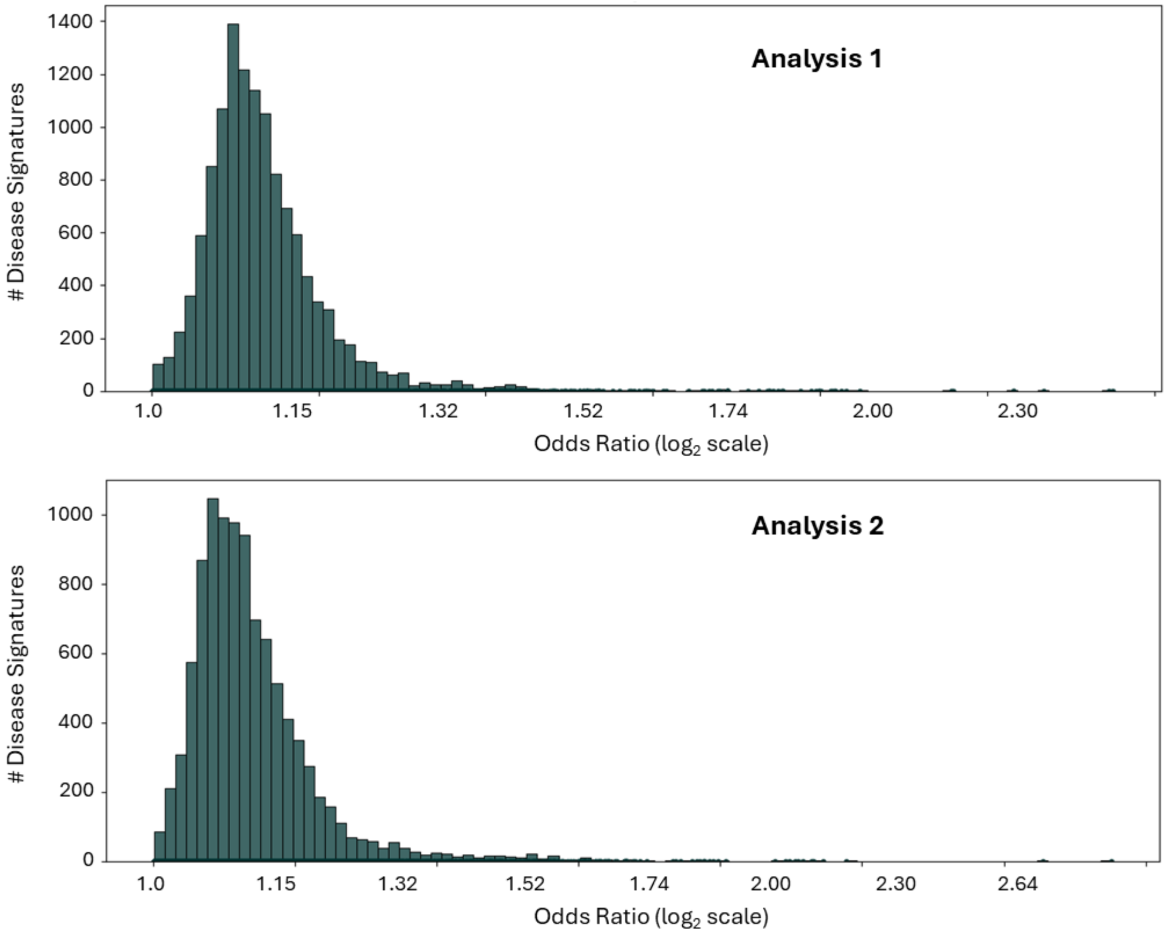


**Supplementary Figure 4.** Distribution of odds ratios of double-refined signatures. a) Odds ratios of double-refined signatures from Analysis 1 in Analysis 1 DecodeME Refinement dataset (Cohort C+G). b) Odds ratios of double-refined signatures from Analysis 2 in Analysis 2 DecodeME Refinement dataset (Cohort C+F).

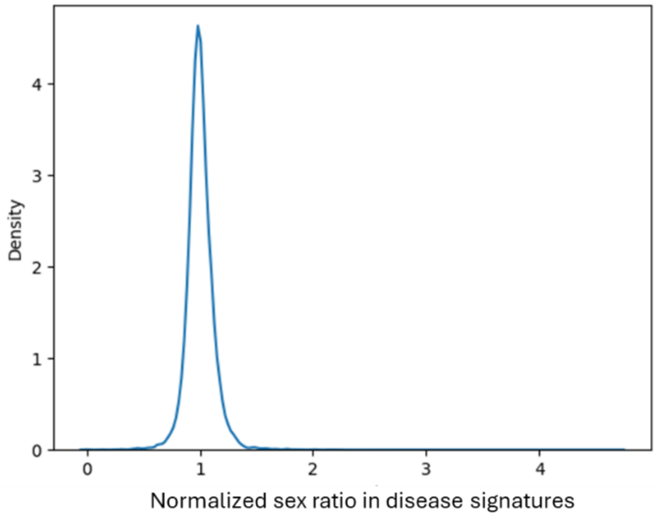


**Supplementary Figure 5.** Density plot of the sex ratio (#males : #females) for each disease signature normalized by the total male and female counts across 20,496 disease signatures. Ratio=1 indicates that the signature occurs at equal frequencies in both sexes.

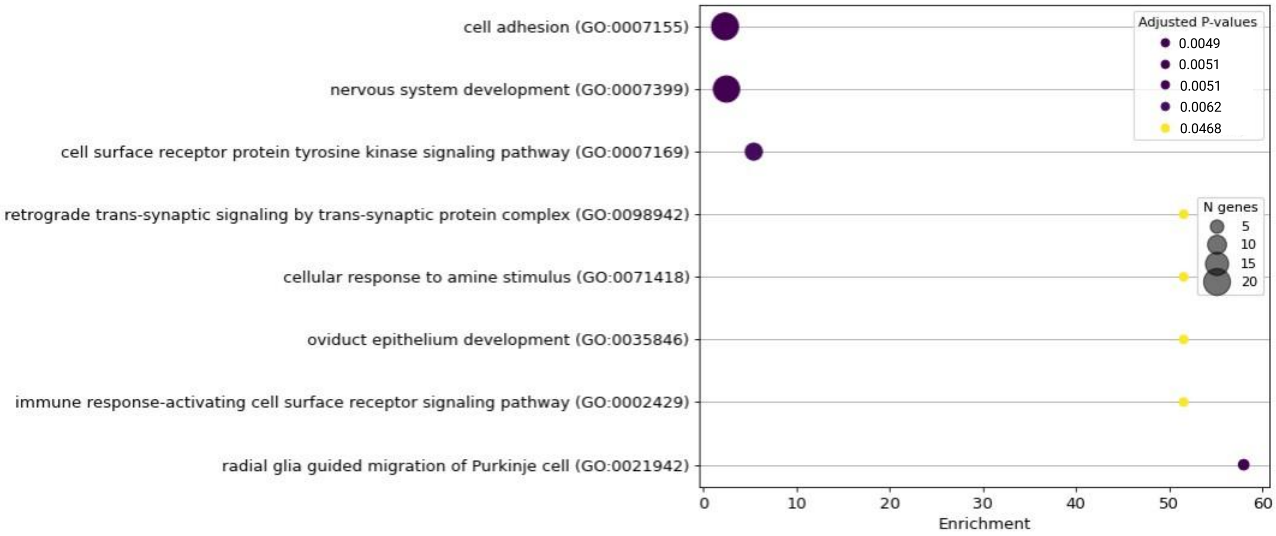


(a)

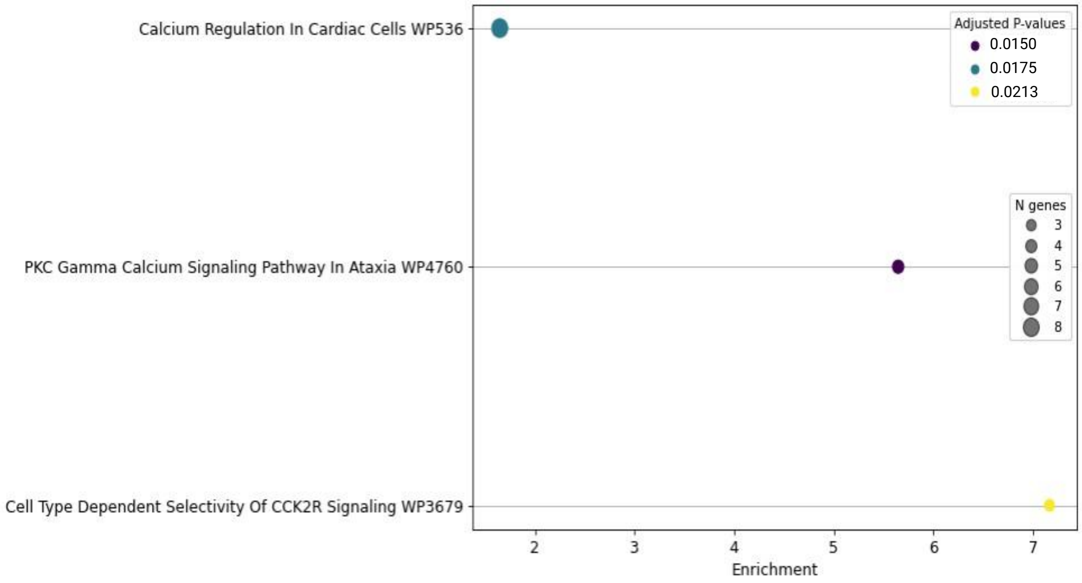


(b)

**Supplementary Figure 6.** Gene set enrichment plot for significant biological processes using (a) Gene Ontology and (b) WikiPathways for the core gene set (n=259) mapped to combinatorial disease signatures. In the dotplots, the x-axis shows enrichment score values, i.e., how strongly a gene set is overrepresented for each biological process, the dot size indicates the number of genes found each process, and the colors reflect statistical significance (adjusted p-values < 0.05).

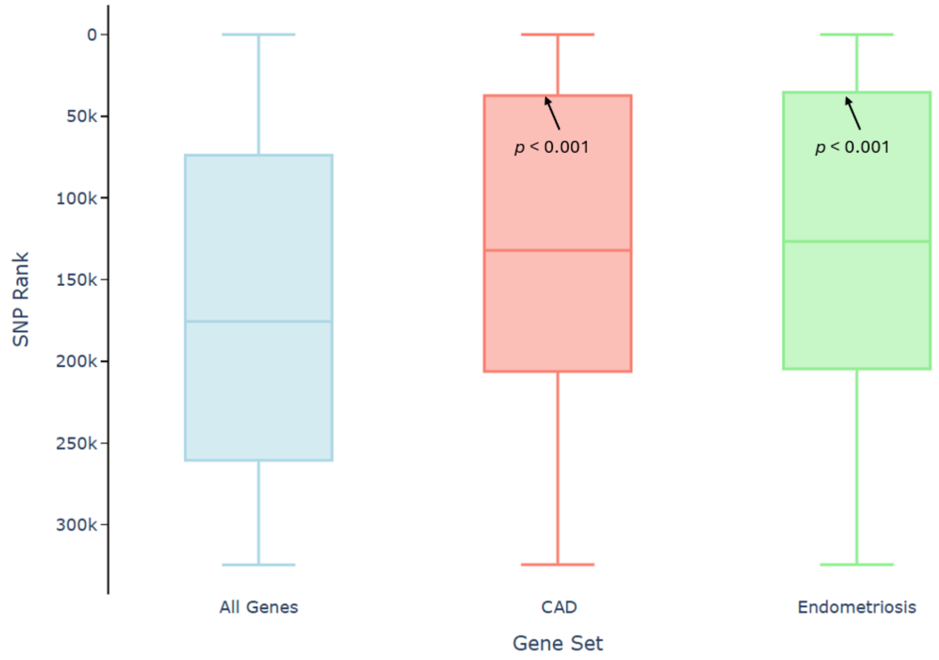


**Supplementary Figure 7.** SNP ranks in the ML model for ME for all SNPs assigned to autosomal protein-coding genes (left) and SNPs assigned to genes associated with coronary artery disease (CAD) and endometriosis in UKB. Higher SNP ranks (i.e. higher positions on the y-axis) denote high feature importance in the ML model. Boxes represent the top 25th and bottom 25th percentile ranks for SNPs assigned to each gene set with solid line denoting median SNP rank. p-values are the probability of randomly observing a top 25th percentile SNP rank that high based on 10,000 gene subsets of equivalent size randomly sampled from the full list of autosomal protein-coding genes.

### Supplementary Tables

**Supplementary Table 1.** UK Biobank control selection criteria for the study.

| Data Type | ICD-10 / UKB Data Field ID | Description |
|---|---|---|
| HES / primary care | G93.3 | Postviral fatigue syndrome |
| HES / primary care | R53.x | Malaise and fatigue |
| HES / primary care | G47.x | Sleep disorders |
| HES / primary care | M79.7 | Fibromyalgia |
| Self-reported | 20002 | Chronic fatigue syndrome |
| Self-reported | 120009 | Ever had fibromyalgia syndrome? |
| Self-reported | 120010 | Ever had Chronic Fatigue Syndrome or Myalgic Encephalitis? |
| Self-reported | 120040 | Fatigue severity over the past week = severe/moderate |
| Self-reported | 120114 | Persistent or recurrent tiredness, weariness, or fatigue > 6 months |
| Self-reported | 120120 | Exercise brings on fatigue |
| Self-reported | 120122 | Fatigue interferes with physical functioning |
| Self-reported | 120123 | Fatigue causes frequent problems |
| Self-reported | 120124 | Fatigue prevents sustained physical functioning |
| Self-reported | 120125 | Fatigue interferes with carrying out certain duties or responsibilities |
| Self-reported | 120126 | Fatigue is among three most disabling symptoms |
| Self-reported | 120127 | Fatigue interferes with work, family or social life |
| Primary care | Xa01F | Chronic fatigue syndrome Myalgic encephalomyelitis ME Myalgic encephalomyelitis Myalgic encephalomyelitis syndrome Postviral fatigue syndrome PVFS - Postviral fatigue syndrome CFS |
| Primary care | F286. | Chronic fatigue syndrome Myalgic encephalomyelitis Myalgic encephalomyelitis ME - Myalgic encephalomyelitis Myalgic encephalomyelitis syndrome Postviral fatigue syndrome PVFS - Postviral fatigue syndrome PVFS - Postviral fatigue syndrome CFS - Chronic fatigue syndrome |
| Primary care | F2860 | Mild chronic fatigue syndrome |
| Primary care | F2861 | Moderate chronic fatigue syndrome |
| Primary care | F2862 | Severe chronic fatigue syndrome |
| Primary care | XaPom | Mild chronic fatigue syndrome |
| Primary care | XaPon | Moderate chronic fatigue syndrome |
| Primary care | XaPoo | Severe chronic fatigue syndrome |
| Primary care | XaPeC | Activity management for chronic fatigue syndrome Activity management for myalgic encephalopathy |

| | | |
|---|---|---|
| Primary care | 8Q1.. | Activity management for chronic fatigue syndrome Activity management for myalgic encephalopathy |
| Primary care | XaR7C | Referral to chronic fatigue syndrome specialist team Referral to myalgic encephalomyelitis specialist team |
| Primary care | XaRAz | Referral for chronic fatigue syndrome activity management Referral for myalgic encephalopathy activity management |
| Primary care | 8HkW. | Referral to chronic fatigue syndrome specialist team Referral to myalgic encephalomyelitis specialist team |

**Supplementary Table 2.** Number of female and male cases in DecodeME cohorts used in this study, broken down by batch.

| ME Cohort | # Total cases | # Female Cases (%) | # Male Cases (%) |
|---|---|---|---|
| DecodeME Batch 1 (Cohort F) | 3405 | 2767 (81.3%) | 638 (18.7%) |
| DecodeME Batch 2 total | 4349 | 3685 (84.7%) | 664 (15.8%) |
| DecodeME Batch 3 total | 4238 | 3595 (84.8%) | 643 (15.2%) |
| DecodeME Batch 2+3 Split 1 (Cohort G) | 3585 | 3032 (84.6%) | 553 (15.4%) |
| DecodeME Batch 2+3 Split 2 (Test) (Cohort H) | 3579 | 3031 (84.7%) | 548 (15.3%) |

**Supplementary Table 3.** Table showing the number of significant SNP networks and core genes count at different case prevalence thresholds. In this study, a case prevalence threshold of 20% was selected for biological interpretation and pathway enrichment analysis.

| Case Prevalence Threshold (%) | No. of significant SNP networks | Core gene count |
|---|---|---|
| > 10% | 1694 | 330 |
| > 20% | 1352 | 259 |
| > 30% | 1050 | 148 |
| > 40% | 745 | 50 |

**Supplementary Table 4.** Public data sources used for annotating SNPs and genes in the PrecisionLife platform.

| Annotation Type | Data Sources |
|---|---|
| Genomic Context | Ensembl[1], Entrez[2], gnomAD[3], HaploReg[4] |
| Encoded Protein Structure and Function | Uniprot[5], InterPro[6], PDBe-KB[7] |
| SNP-Disease or Trait Association | dbSNP[8], GWAS Catalog[9], PharmGKB[10], Open Targets[11] |
| Gene-Disease or Trait Association | Open Targets[11], PHAROS[12], Mouse Genome Informatics[13], Human Phenotype Ontology[14], OMIM[15] |
| Gene Tissue/Cell Expression | Human Protein Atlas[16], GTEx[17], Expression Atlas[18] |
| Pathways or MoA | Gene Ontology[19], Reactome[20], WikiPathways[21] |
| Drugs and Chemical Compounds | Global Data[22], ChEMBL[23], DrugBank[24], PubChem[25] |

**Supplementary Table 11.** Case/control criteria used to identify UKB cohort for combinatorial analysis of coronary artery disease.

**CAD Case Inclusion Criteria**

1. Individuals who were between 40-69 years at the time of UKB recruitment (Datafield 21022)
2. CAD diagnosis must satisfy any one of the following:
3. Individuals with any of the following codes:
    a. ICD-10 codes in Hospital Episode Statistics (HES, Datafield 41270)
        i. I21 (acute myocardial infarction)
        ii. I22 (subsequent myocardial infarction)
        iii. I23 (certain current complications following acute myocardial infarction)
        iv. I24.1 (Dressler's syndrome)
        v. I25.2 (old myocardial infarction)
    b. OPCS4 codes in Operative procedures (Datafield 41272)
        i. K40.* (Saphenous vein graft replacement of coronary artery)
        ii. K41.* (Other autograft replacement of coronary artery)
        iii. K45.* (Connection of thoracic artery to coronary artery)
        iv. K49.1 (Percutaneous transluminal balloon angioplasty of one coronary artery)
        v. K49.2 (Percutaneous transluminal balloon angioplasty of multiple coronary artery)
        vi. K49.8 (Other specified transluminal balloon angioplasty of coronary artery)
        vii. K49.9 (Unspecified transluminal balloon angioplasty of coronary artery)
        viii. K50.2 (Percutaneous transluminal coronary thrombolysis using streptokinase)
        ix. K75.* (Percutaneous transluminal balloon angioplasty and insertion of stent into coronary artery)
    c. Self-reported codes
        i. 1075 (heart attack/myocardial infarction) in Non-cancer illness code (Datafield 20002)
        ii. Self-report of heart attack or myocardial infarction in Vascular/heart problems diagnosed by doctor (Datafield 6150)
    d. Self-reported Operation codes (Datafield 20004)
        i. 1070 (coronary angioplasty (ptca) +/- stent)
        ii. 1095 (coronary artery bypass grafts (cabg))

**CAD Controls Inclusion / Exclusion Criteria**

*Inclusion criteria:*

1. Gender-matched, oldest possible individuals
2. Individuals who were between 40-69 years at the time of UKB recruitment (Datafield 21022)
3. European genetic ancestry

*Exclusion criteria:*

1. Remove individuals that meet any of the above criteria for case diagnosis
2. Remove individuals with family history of heart disease reported in:
    a. Illnesses of mother (Datafield 20110)
    b. Illnesses of father (Datafield 20107)
    c. Illnesses of siblings (Datafield 20111)

**Supplementary Table 6.** Number of signatures and component SNPs from the initial DecodeME combinatorial analyses, after Refinement in a UKB cohort, and after further Refinement in an independent DecodeME cohort. Core genes were selected based on association with ME and case prevalence in a DecodeME Refinement dataset.

| Analysis | # Initial signatures (# SNPs) | # UKB-refined signatures (# SNPs) | # Double-refined signatures (# SNPs) | # Genes mapping to double-refined signatures |
|---|---|---|---|---|
| Analysis 1 | 150,683 (4,209) | 43,816 (3,679) | 12,463 (2,915) | 1,001 |
| Analysis 2 | 95,916 (8,101) | 31,612 (7,229) | 9,963 (5,008) | 1,647 |
| Combined Results | -- | -- | 22,411 (7,555) | 2,311 |

**Supplementary Table 7.** Number of SNPs in combinatorial disease signatures identified in this study.

| No. of SNPs in Combination | No. of Double-Refined Signatures |
|---|---|
| 1 | 610 |
| 2 | 13,117 |
| 3 | 7,223 |
| 4 | 1,461 |
| **Total** | **22,411** |

**Supplementary Table 8.** Gene overlap (n=14) between disease signatures identified in this study and DecodeME 2025 study[26] (including both Tier 1, Tier 2 and the gene linked to chronic pain). Five Tier 1 lead genes that map to the 8 GWAS significant loci reported by DecodeME are shown in bold. All genomic positions in the table are in GRCh37.

| Gene | DecodeME Association | Locus | Disease signature count for SNP |
|---|---|---|---|
| ***BTN2A2*** | **GWAS-1**- Tier 1 lead | *6p22.2* | 33 |
| ***CA10*** | **GWAS-1**- Tier 1 lead | *17q22* | 1 |
| ***CA10*** | **GWAS-1**- Tier 1 lead | chr17q22 | 2 |
| ***CA10*** | **GWAS-1**- Tier 1 lead | chr17q22 | 1 |
| ***CSE1L*** | **GWAS-1** Tier 1 lead | chr20q13.13 | 160 |
| ***OLFM4*** | **GWAS-Infection**- Tier 1 lead | *13q14.3* | 7 |
| ***SUDS3*** | **GWAS-2**- Tier 1 lead | chr17q24.23 | 116 |
| *ABT1* | GWAS-1 - Tier 1 other | *6p22.2* | 44 |
| *DARS2* | GWAS-1 – Tier 1 other | chr1q25.1 | 14 |
| *DCC* | Other - chronic pain | *18q21.2* | 10 |
| *DCC* | Other - chronic pain | *18q21.2* | 30 |
| *DCC* | Other - chronic pain | *18q21.2* | 132 |
| *DCC* | Other - chronic pain | *18q21.2* | 55 |
| *DCC* | Other - chronic pain | *18q21.2* | 55 |

| | | | |
|---|---|---|---|
| *DCC* | Other - chronic pain | chr18q21.2 | 2 |
| *DCC* | Other - chronic pain | chr18q21.2 | 1 |
| *DCC* | Other - chronic pain | chr18q21.2 | 1 |
| *DCC* | Other - chronic pain | chr18q21.2 | 1 |
| *DDX27* | GWAS-1 Tier 1 other | chr20q13.13 | 67 |
| *HFE* | GWAS-1 - Tier 1 other | chr6p22.2 | 3 |
| *TRIM38* | GWAS-1 - Tier 1 other | chr6p22.2 | 2 |
| *TRIM38* | GWAS-1 - Tier 1 other | chr6p22.2 | 5 |
| *VSIG10* | GWAS-2- Tier 1 other | chr12q24.23 | 6 |
| *ZNFX1* | GWAS-1 Tier 1 other | chr20q13.13 | 67 |
| *ZNF322* | GWAS-1 - Tier 1 other | chr6p22.2 | 4 |
| *ZNF322* | GWAS-1 - Tier 1 other | chr6p22.2 | 6 |

**Supplementary Table 9.** List of 102 genes identified in double-refined signatures for ME and previous Sano GOLD long COVID study (Severe, Fatigue Dominant or General sub cohort analyses). 76 of these genes were reproduced in the All of Us long COVID cohort[Error! Reference source not found.,28].

| Gene | Long COVID cohort in Sano Genetics GOLD study | Reproduced in All of Us long COVID cohort |
|---|---|---|
| *ADAM12* | General | Yes |
| *ADAMTS9* | General | Yes |
| *ADGRL2* | General | Yes |
| *ANKS1B* | General | Yes |
| *ANO4* | General | Yes |
| *ASIC2* | General | Yes |
| *CACNA2D3* | General | Yes |
| *CACNB2* | General | Yes |
| *CAMTA1* | General | Yes |
| *CDH12* | General | Yes |
| *CDH13* | General | Yes |
| *CDK14* | Fatigue Dominant | Yes |
| *CGNL1* | General | Yes |
| *CLCN6* | Severe | Yes |
| *CPVL* | General | Yes |
| *CREB5* | General | Yes |
| *CSGALNACT1* | General | Yes |
| *CSMD2* | General | Yes |
| *DENND1A* | General | Yes |
| *DISC1* | General | Yes |
| *DPP6* | General | Yes |
| *ERBB4* | General | Yes |

| | | |
|---|---|---|
| *EYA1* | General | Yes |
| *F13A1* | General | Yes |
| *FGF12* | General | Yes |
| *GABBR2* | General | Yes |
| *GUCY1A2* | Severe, Fatigue Dominant | Yes |
| *HMGCLL1* | General | Yes |
| *HPN* | Fatigue Dominant | Yes |
| *JAKMIP2* | Fatigue Dominant, General | Yes |
| *KCNMA1* | General | Yes |
| *KIF26B* | General | Yes |
| *KRT86* | General | Yes |
| *MAPK9* | Fatigue Dominant | Yes |
| *MSR1* | General | Yes |
| *MTMR7* | General | Yes |
| *MYO3B* | Severe | Yes |
| *NELL1* | General | Yes |
| *OPCML* | General | Yes |
| *PACRG* | General | Yes |
| *PCDH9* | Severe | Yes |
| *PDZD2* | General | Yes |
| *POR* | Fatigue Dominant | Yes |
| *PPP1R16B* | Severe | Yes |
| *PRODH* | Severe | Yes |
| *RBFOX1* | Severe | Yes |
| *RIMS2* | General | Yes |
| *RRBP1* | Fatigue Dominant | Yes |
| *RYR2* | General | Yes |
| *RYR3* | General | Yes |
| *SCUBE1* | General | Yes |
| *SEL1L* | General | Yes |
| *SLC25A21* | General | Yes |
| *SLC38A4* | General | Yes |
| *SMARCA2* | General | Yes |
| *SNTG1* | General | Yes |
| *SNX31* | General | Yes |
| *SORCS2* | Severe | Yes |
| *SOX5* | Severe | Yes |
| *ST8SIA1* | General | Yes |
| *SUCLA2* | General | Yes |
| *SYN3* | Fatigue Dominant | Yes |
| *TACR1* | General | Yes |

| | | |
|---|---|---|
| *TBXAS1* | General | Yes |
| *TENM3* | Severe | Yes |
| *TNIK* | Fatigue Dominant | Yes |
| *TRIM9* | General | Yes |
| *TRPM3* | General | Yes |
| *TSPAN5* | General | Yes |
| *TTC39C* | General | Yes |
| *USH2A* | General | Yes |
| *USP30* | General | Yes |
| *XKR3* | General | Yes |
| *ZNF385D* | General | Yes |
| *ZNF577* | General | Yes |
| *ZNF71* | General | Yes |
| *ADAM28* | General | |
| *ADIPOQ* | Severe | |
| *CCDC149* | Severe | |
| *CNTN4* | Fatigue Dominant | |
| *COL27A1* | General | |
| *DLC1* | Severe | |
| *DSCAML1* | Severe | |
| *FAM53B* | General | |
| *KLF12* | Fatigue Dominant | |
| *MAML2* | General | |
| *NAV3* | General | |
| *NLGN1* | Fatigue Dominant | |
| *PCSK2* | Severe, Fatigue Dominant | |
| *PITPNC1* | Fatigue Dominant | |
| *PLXNA2* | Fatigue Dominant | |
| *PRKG1* | General | |
| *PSMB9* | General | |
| *PTPRD* | Fatigue Dominant | |
| *RADIL* | General | |
| *SPTBN5* | Fatigue Dominant | |
| *SYNPR* | General | |
| *TAP1* | General | |
| *TIAM2* | General | |
| *TNS1* | Fatigue Dominant | |
| *WWOX* | Fatigue Dominant | |
| *ZMIZ1* | Severe | |

**Supplementary Table 10.** Results of length-matched permutation test used to assess enrichment of long COVID genes across three gene length bins for 2311 genes mapping to double-refined ME signatures. Enrichment in all gene length bins is significant ($p < 0.017$) after Bonferroni multiple-testing correction (shown in bold).

| Gene length bins (kb) | Background gene count | ME gene count (n=2311) | Overlap of long COVID genes (n=102) | Mean expected overlap across 10,000 random permutations | *p* value |
|---:|---:|---:|---:|---:|---:|
| <= 50 | 13723 | 692 | 10 | 2.20 | **0.0002** |
| 50 - 250 | 5579 | 1061 | 33 | 18.23 | **0.0004** |
| > 250 | 1034 | 558 | 59 | 43.21 | **0.0003** |

**Supplementary Table 11.** Overlap between genes associated with long COVID and the 97 'top-ranked' genes from a machine learning model for ME (i.e., genes that map to the top 20 ranked SNPs in at least one fold).

| Long COVID phenotype | # of Long COVID genes | # Genes overlapping with ML model for ME | *p*-value for enrichment | Overlapping genes |
|---|---:|---:|---:|---:|
| Fatigue-Dominant | 34 | 3 | 0.001 | *MAPK9*, *NLGN1*, *PTPRD* |
| Severe | 38 | 1 | 0.18 | *GPC6* |
| General | 153 | 2 | 0.19 | *MYOCD*, *OPCML* |
